# Assessment of valvular function in over 47,000 people using deep learning-based flow measurements

**DOI:** 10.1101/2023.04.29.23289299

**Authors:** Shinwan Kany, Joel T. Rämö, Cody Hou, Sean J. Jurgens, Victor Nauffal, Jon Cunningham, Emily S. Lau, Atul J. Butte, Jennifer E. Ho, Jeffrey E. Olgin, Sammy Elmariah, Mark E. Lindsay, Patrick T. Ellinor, James P. Pirruccello

**Author notes:** Corresponding author: James P. Pirruccello, MD; Division of Cardiology, University of California San Francisco, 555 Mission Bay Blvd South, Box 3118, San Francisco CA 94158.

## Abstract

Valvular heart disease is associated with a high global burden of disease. Even mild aortic stenosis confers increased morbidity and mortality, prompting interest in understanding normal variation in valvular function at scale.

We developed a deep learning model to study velocity-encoded magnetic resonance imaging in 47,223 UK Biobank participants. We calculated eight traits, including peak velocity, mean gradient, aortic valve area, forward stroke volume, mitral and aortic regurgitant volume, greatest average velocity, and ascending aortic diameter. We then computed sex-stratified reference ranges for these phenotypes in up to 31,909 healthy individuals. In healthy individuals, we found an annual decrement of 0.03cm^2^ in the aortic valve area. Participants with mitral valve prolapse had a 1 standard deviation [SD] higher mitral regurgitant volume (P=9.6 × 10^−12^), and those with aortic stenosis had a 4.5 SD-higher mean gradient (P=1.5 × 10^−431^), validating the derived phenotypes’ associations with clinical disease. Greater levels of ApoB, triglycerides, and Lp(a) assayed nearly 10 years prior to imaging were associated with higher gradients across the aortic valve. Metabolomic profiles revealed that increased glycoprotein acetyls were also associated with an increased aortic valve mean gradient (0.92 SD, P=2.1 x 10^−22^). Finally, velocity-derived phenotypes were risk markers for aortic and mitral valve surgery even at thresholds below what is considered relevant disease currently.

Using machine learning to quantify the rich phenotypic data of the UK Biobank, we report the largest assessment of valvular function and cardiovascular disease in the general population.

## Introduction

Valvular heart disease is associated with mortality and a high global burden of disease; in 2017, over 30 million people were living with aortic or mitral valve disease ^1^. Consequently, there has been considerable interest in studying the natural history of severe valvular disease ^2^. More recently, mild-to-moderate aortic stenosis has been shown to portend increased risk of cardiovascular events or death ^3–5^. These observations motivate the analysis of normal variation in valvular function within a generally healthy population and a reassessment of the clinical consequences of valvular abnormalities below established disease thresholds. The failure of pharmacologic trials for established aortic stenosis also adds urgency to efforts to gain more insight into measurements of valvular function prior to the onset of clinically apparent disease ^6, 7^.

Valvular pathologies such as aortic stenosis (AS) and mitral regurgitation have anatomic characteristics that can be visualized, but functional assessment using pressure gradients is needed to understand disease severity ^8^. Echocardiography—which is non-invasive and low cost—is the standard imaging modality recommended by clinical guidelines to assess hemodynamic effects of valvular heart disease. Cardiovascular magnetic resonance imaging (cMRI) is another noninvasive imaging modality that can assess cardiac and valvular function, offering lower intra- and interobserver variability compared with echocardiography ^9^. Furthermore, phase-contrast cMRI can provide blood flow velocity information and aortic flow patterns to assess valvular and aortic disease ^10^. Cost and high barriers for access to cMRI have prevented large scale assessments of valvular heart disease so far. Recently, deep learning models have been developed to measure anatomical structures with cMRI such as aortic diameter at scale in large biobanks like the UK Biobank ^11, 12^. These big-data approaches have also enabled the development of new clinical risk prediction models ^13^. The velocity-encoded images in the UK Biobank have been previously used to calculate aortic valve area (AVA) by planimetry in a supravalvular level and for greatest average velocity in the ascending aorta ^14, 15^. However, to date, a large-scale analysis of clinically relevant measures of valvular function has not been reported.

Anchored by a deep learning model that isolates aortic flow from velocity-encoded cMRI, this study aimed to assess aortic and mitral valve measurements and to evaluate their associations with clinical risk factors and disease phenotypes in nearly 50,000 UK Biobank participants.

## Results

### Deriving valvular flow-based phenotypes with deep learning

We hypothesized that the phase contrast imaging sequences in the cardiac MRI dataset of the UK Biobank, captured just above the aortic valve, could be used to accurately quantify velocities in the ascending aorta, enabling assessment of flow across the aortic valve (Figure 1). One cardiology fellow (SK) manually annotated 950 images that were randomly selected from the UK Biobank imaging series of velocity encoded sequences (Figure 2). A PyTorch-based U-Net model was trained using 800 images in the training set and 150 in the validation set (**Online Methods**) ^16^. In 30 separate images annotated by a board-certified cardiologist (JPP), the model achieved a mean Dice score of 0.964 (95% CI 0.957 to 0.971) for the ascending aortic blood pool.

**Figure 1:**
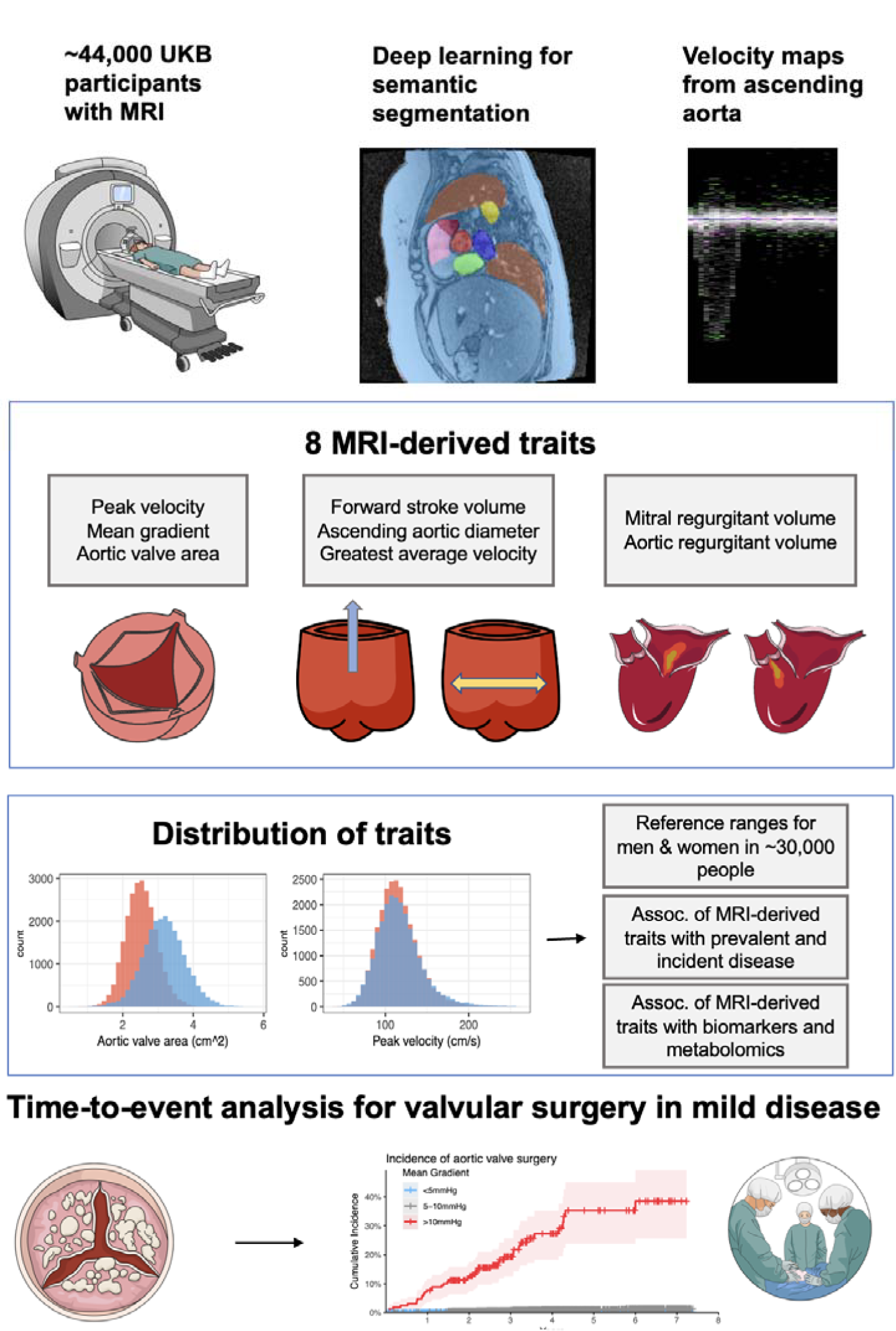
Study overview. Study overview: Deep learning was used to create segmentation masks of the ascending aorta in the aortic flow image series in the UK Biobank. The masks were used to create velocity maps that were used to construct 8 MRI-derived phenotypes. For mitral regurgitant volume the left ventricular stroke volume was calculated using left ventricular end-diastolic and end-systolic volume. The traits were then used to create reference ranges stratified by sex and age in healthy individuals. Logistic regression and Cox hazard models were calculated for the association with prevalent and incident disease. Additionally, linear regressions were performed for the association of the traits with biomarkers and metabolomic taken ∼9.6 years before MRI. Finally, we analyzed time to incident valve surgery based on different thresholds for mild disease. Medical images were used from Servier Medical Art and Mind the Graph under Creative Commons 3.0 license.

**Figure 2:**
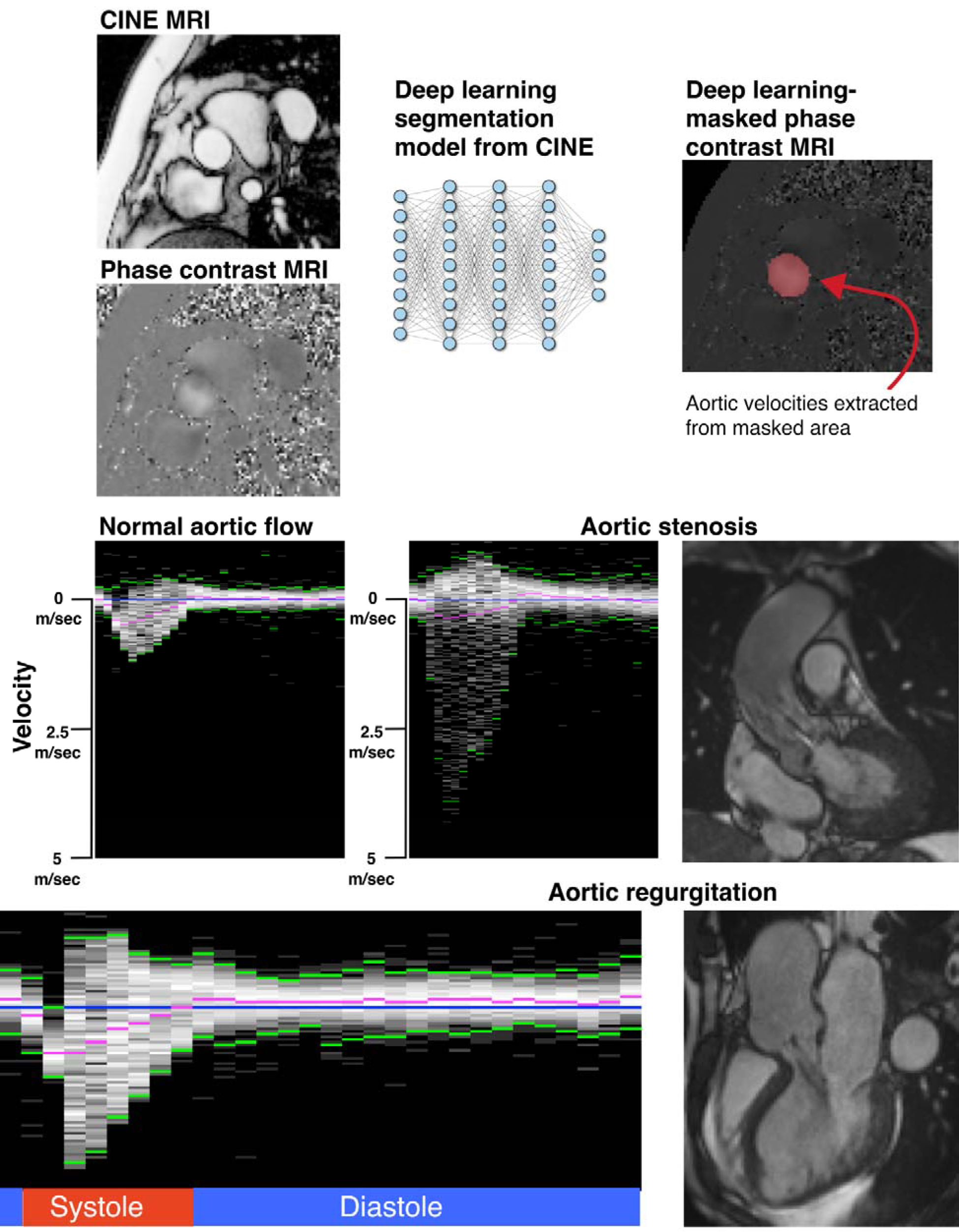
Graphical overview of workflow. Overview of workflow: Images from CINE series were ingested for semantic segmentation to create masks allowing extraction of velocity-encoded pixels corresponding to the ascending aorta. In post processing, the cardiac cycle was reconstructed (30 images/sec) to determine the flow measurements corresponding to systole and diastole and to calculate all measurements. Exemplary velocity tracings and corresponding anatomical MRI images shown for normal aortic flow, aortic stenosis and aortic regurgitation. In the velocity tracings, velocity increases towards the bottom of the image; the blue line represents the zero-point for velocity; the magenta points represent the mean velocity; and the green points represent the 1^st^ and 99^th^ percentile velocities. The anatomical image shown for the aortic stenosis panel is a view of the left ventricular outflow tract during systole where a jet can be seen over the aortic valve. The anatomical image for the aortic regurgitation panel is a 3-chamber view during diastole where aortic regurgitant flow can be seen impinging on the anterior mitral leaflet. For the regurgitant flow panel, net negative mean flow can be seen throughout diastole. Cardiac images are reproduced by kind permission of UK Biobank ©.

A total of 49,798 individuals were eligible to contribute to analyses **(Supplementary Figure AA**). The model was applied to all participants, and results were post-processed and quality-controlled (exclusion of N=2,576; **Online Methods**) to produce eight phenotypes in 47,223 participants (Table 1) who contributed to at least one trait (Figure 1). An overview of the phenotypes and their distributions is presented in **Supplementary Figure BB**.

**Table 1:**
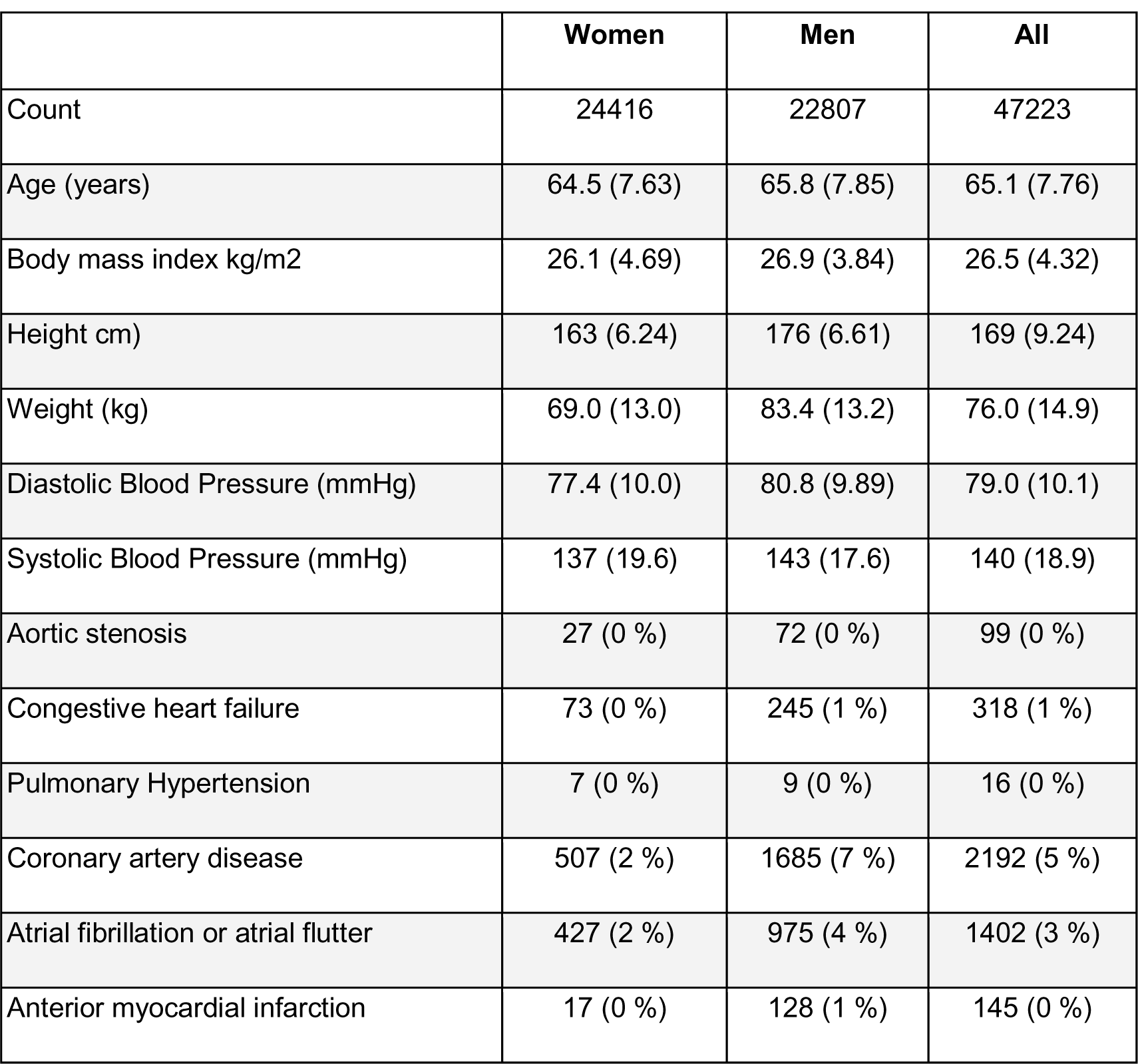
Participants characteristics. Clinical characteristics of the 47,223 participants at the time of MRI whose data contributed to at least one phenotype. For quantitative phenotypes, values shown represent mean (s.d.). For count data, values shown represent count (%).

|  | Women | Men | All |
| --- | --- | --- | --- |
| Count | 24416 | 22807 | 47223 |
| Age (years) | 64.5 (7.63) | 65.8 (7.85) | 65.1 (7.76) |
| Body mass index kg/m2 | 26.1 (4.69) | 26.9 (3.84) | 26.5 (4.32) |
| Height cm) | 163 (6.24) | 176 (6.61) | 169 (9.24) |
| Weight (kg) | 69.0 (13.0) | 83.4 (13.2) | 76.0 (14.9) |
| Diastolic Blood Pressure (mmHg) | 77.4 (10.0) | 80.8 (9.89) | 79.0 (10.1) |
| Systolic Blood Pressure (mmHg) | 137 (19.6) | 143 (17.6) | 140 (18.9) |
| Aortic stenosis | 27 (0 %) | 72 (0 %) | 99 (0 %) |
| Congestive heart failure | 73 (0 %) | 245 (1 %) | 318 (1 %) |
| Pulmonary Hypertension | 7 (0 %) | 9 (0 %) | 16 (0 %) |
| Coronary artery disease | 507 (2 %) | 1685 (7 %) | 2192 (5 %) |
| Atrial fibrillation or atrial flutter | 427 (2 %) | 975 (4 %) | 1402 (3 %) |
| Anterior myocardial infarction | 17 (0 %) | 128 (1 %) | 145 (0 %) |

Of the three measures of flow limitation across the aortic valve (aortic valve area [AVA], peak velocity, and mean gradient), the peak velocity and mean gradient measures were closely correlated (r=0.92); they had modest inverse correlations with AVA (r=-0.45 and r=-0.39, respectively) (**Supplementary Figure CC**). The AVA had a similar degree of correlation with ascending aortic diameter (r=0.48); in contrast, the correlation between peak velocity and ascending aortic diameter was r=-0.1. Aortic regurgitant volume (r=0.36) and mitral regurgitant volume (r=0.26) were also positively correlated with aortic diameter.

### Valvular function over 30,000 healthy people

After excluding participants with diagnostic codes consistent with a history of AS, coronary artery disease, heart failure, or hypertension, valvular phenotype distributions were calculated by groups of age and sex in up to 31,909 participants (**Supplementary Table C and Supplementary Figure DD**). Reference ranges for the valvular phenotypes were defined for men and women using the observed normal ranges across age groups (Table 2). AVA was lower in older age groups, whereas peak velocity and mean gradient remained similar across age groups. Compared to women, men had greater AVA but similar peak velocity and mean gradient estimates. Formalized in a statistical model, age and sex explained virtually none of the variation in peak velocity or mean gradient (model R^2^ 0.001 and 0.004, respectively), while AVA was the most strongly associated with age and sex of all tested phenotypes (model R^2^ 0.35; **Supplementary Table C**). On average, after accounting for age, men had a 1.1 standard deviation (SD) greater AVA than women (0.69cm^2^ area difference, P=3.4 × 10^−2760^ against a null hypothesis of no difference), while the difference for peak velocity was less than 1cm/sec (P=1.8 × 10^−2^). The observed sex difference in AVA was only partially accounted for by indexing on body surface area (effect of male sex 0.56 SD, P=5.0 × 10^−562^).

**Table 2:** Reference ranges for the cardiac phenotypes in healthy individuals. Reference ranges calculated and stratified by sex in up to 31,909 participants. Participants with a known history of AS, coronary artery disease, heart failure, or hypertension were excluded. For each phenotype, five zones were defined: abnormally low (below the 5% cutoff in all age groups, orange), borderline low (below the 5% cutoff in at least one age group, yellow), normal (between the 5% and 95% cutoff in all age groups), borderline high (above the 95% cutoff in at least one age group, yellow), and abnormally high (above the 95% cutoff in all age groups, orange).

|  | Men |  |  |  |  |
| --- | --- | --- | --- | --- | --- |
| Trait | Abnormal low |  | Normal |  | Abnormal high |
| Aortic valve area (cm<sup>2</sup>) | 2.1 |  | 2.5 - 3.9 |  | 4.4 |
| Aortic regurgitant volume (mL) | 1.7 |  | 1.9 - 12.7 |  | 16.4 |
| Mitral regurgitant volume (mL) | 2.4 |  | 2.6 - 29.7 |  | 30.5 |
| Forward stroke volume (mL) | 50 |  | 61 - 100 |  | 114 |
| Ascending aortic diameter (cm) | 2.9 |  | 3.0 - 4.0 |  | 4.1 |
| Peak velocity (cm/sec) | 73 |  | 81 - 155 |  | 163 |
| Greatest average velocity (cm/sec) | 27 |  | 36 - 59 |  | 76 |
| Mean gradient (mmHg) | 1.2 |  | 1.5 - 4.6 |  | 4.9 |
|  | Women |  |  |  |  |
| Aortic valve area (cm<sup>2</sup>) | 1.7 |  | 2.0 - 3.1 |  | 3.4 |
| Aortic regurgitant volume (mL) | 0.8 |  | 0.9 - 8.9 |  | 10.4 |
| Mitral regurgitant volume (mL) | 2.0 |  | 2.3 - 25.1 |  | 25.7 |
| Forward stroke volume (mL) | 42 |  | 51 - 79 |  | 92 |
| Ascending aortic diameter (cm) | 2.7 |  | 2.8 - 3.6 |  | 3.7 |
| Peak velocity (cm/sec) | 75 |  | 83 - 153 |  | 166 |
| Greatest average velocity (cm/sec) | 26 |  | 34 - 55 |  | 69 |
| Mean gradient (mmHg) | 1.2 |  | 1.5 - 4.5 |  | 4.9 |

Age and sex explained 28% of the variance in forward stroke volume, with a 4.6mL lower stroke volume (−0.3 SD, P=1.0 × 10^−436^) per decade. Men had a 16.3mL greater stroke volume than women (1.0 SD difference, P=4.4 × 10^−2009^). Aortic regurgitant volume was greater in men (2.4mL, P=2.4 × 10^−688^), but in contrast to forward stroke volume, it was also greater in older participants (0.4mL per decade, P=2.6 × 10^−45^). Mitral regurgitant volume was only weakly explained by age and sex (model R^2^ 0.024), with the observation driven by sex (2.5mL greater in men, P=2.9 × 10^−151^) but not age (P=8.7 × 10^−1^).

Within the same healthy subgroup, repeat measurements were available in up to 2,969 participants who had undergone repeat imaging (mean 2.6 ± 1.0 years after the initial visit). Over that follow-up time, no change over time in peak velocity (P=5.4 × 10^−1^) or mean gradient (P=3.6 × 10^−1^) was observed (**Supplementary Table D**). AVA declined by 0.03cm^2^ (0.05 SD) per year (P=2.3 × 10^−29^ against a null hypothesis of no change). Aortic and mitral regurgitant volumes increased with time (∼0.04 SD per year, P=1.4 × 10^−9^ and P=3.1 × 10^−6^, respectively). However, these models explained little of the variance in the change in phenotypes over time (model R^2^ 0.04 or less).

### Clinical grading of aortic valve function

Using the current recommendations on the assessment of aortic valve stenosis by European and American professional societies ^17^, we graded all participants by three criteria: peak velocity, mean gradient and AVA (**Supplementary Table E**). Participants with severe AS according to clinical criteria were rare: no participant had a grading of severe AS in all three criteria. No participants had severe AS by peak velocity or mean gradient; and 31 by AVA. Similarly, participants with moderate AS were uncommon and ranged from 31 cases (mean gradient) to 295 cases (AVA). Peak velocity is the only trait with a lower limit of what is considered mild disease in the current consensus document by the European Association of Cardiovascular Imaging and the American Society of Echocardiography ^17^, with a range of 2.6 - 2.9 m/s. Only 66 participants fulfilled that definition of mild disease. Keeping the highest severity rating across any of the three measurements for each person, 36 participants had mild disease, 313 had moderate disease, and 31 had severe disease.

### Prevalent cardiovascular disease associated with aortic flow and velocities

To understand which diseases were most strongly associated with the MRI-based phenotypes, we used linear models to test for association between the phenotypes (treated as the independent variable) and cardiovascular diseases diagnosed prior to the time of MRI (**Supplementary Table F**). We began by examining the relationship between each measurement and its canonical disease. As expected, a clinical diagnosis of aortic regurgitation was strongly associated with greater aortic regurgitant volume (N=64, 2.44 SD increase, P=6.3 × 10^−94^). Mean gradient had the largest magnitude of association with the 99 cases of prevalent AS (4.45 SD increase, P=1.4 × 10^−422^). Prevalent AS was also associated with higher peak velocity (3.21 SD, P= 9.9 × 10^−220^) and smaller AVA (-1.69 SD, P=6.6 × 10^−92^). Similarly, the presence of mitral valve prolapse (N=45, 1.02 SD increase, P=9.7 × 10^−12^) or a clinical diagnosis of mitral regurgitation (N=123, 0.61 SD increase, P=6.0 × 10^−11^) was associated with greater mitral regurgitant volume.

We also observed associations between coronary disease risk factors and aortic valve phenotypes. Prevalent hypertension (N=14577) was associated with higher peak velocity (0.29 SD, P= 3.5 × 10^−174^) and greater mean gradient (0.28 SD, P=2.6 × 10^−158^), while the magnitude of its association with smaller AVA (−0.07 SD, P=5.2 × 10^−16^) was more modest. The presence of hypercholesterolemia had a more uniform association with higher peak velocity (0.17 SD, P= 1.3 × 10^−48^), greater mean gradient (0.17 SD, P=5.4 × 10^−47^), and smaller AVA (−0.15 SD, P=5.7 × 10^−56^). Coronary artery disease itself (N=2192) was associated with a higher peak velocity (0.19 SD, P=2.4 × 10^−17^), a greater mean gradient (0.21 SD, P=2.3 × 10^−21^), and a smaller AVA (−0.22 SD, P=7.8 × 10^−32^).

Sample sizes for rare conditions were small, but significant associations were observed for some. For instance, hypertrophic cardiomyopathy was associated with greater mitral regurgitant volume (N=21, 1.2 SD increase, P=6.7 × 10^−08^) and aortic regurgitant volume (N=25, 0.9 SD increase, P=4.0 × 10^−06^). However, there was no significant association between prevalent dilated cardiomyopathy (N=22) and any of the flow-derived phenotypes, likely due to small sample size. A more general diagnosis of heart failure (N=318) was significantly associated with a smaller AVA (−0.24 SD, P=3.4 × 10^−07^).

Examining the non-valve phenotypes (forward stroke volume and greatest average velocity), we observed that greater forward stroke volume was found in the presence of aortic regurgitation (N=64, 0.6 SD, P=1.2 × 10^−07^), obesity (N=1048, 0.38 SD, P=1.5 × 10^−43^), and sleep apnea (N=606, 0.23 SD, P=5.7 × 10^−11^). Chronic obstructive pulmonary disease (N=1098, −0.18 SD, P=1.2 × 10^−11^) and osteoporosis (N=1440, - 0.18 SD, P=4.3 × 10^−14^) were associated with reduced forward stroke volume. For greatest average velocity, a diagnosis of obesity was associated with greater velocity (N=1048, 0.24 SD, P=2.8 × 10^−15^), as were diabetes (N=1860, 0.09 SD, P=3.4 × 10^−05^) and hypertension (N=14576, 0.09 SD, P=1.7 × 10^−20^). Atrial fibrillation (N=1402) was associated with lower greatest average velocity (−0.17 SD, P=2.9 × 10^−11^).

### Relation between valve function and incident cardiovascular diseases

The deep learning model enabled an assessment of the association between the functional valve traits and incident disease outcomes after MRI on a biobank scale. Variation in aortic velocities was associated with several cardiovascular diagnoses associated with one or more MRI-derived phenotype after Bonferroni correction included atrial fibrillation, bradyarrhythmia, CAD, diabetes, endocarditis, heart failure, hypercholesterolemia, hypertension, pulmonary hypertension, and thoracic aortic disease (Figure 3, **Supplementary Table G**).

**Figure 3:**
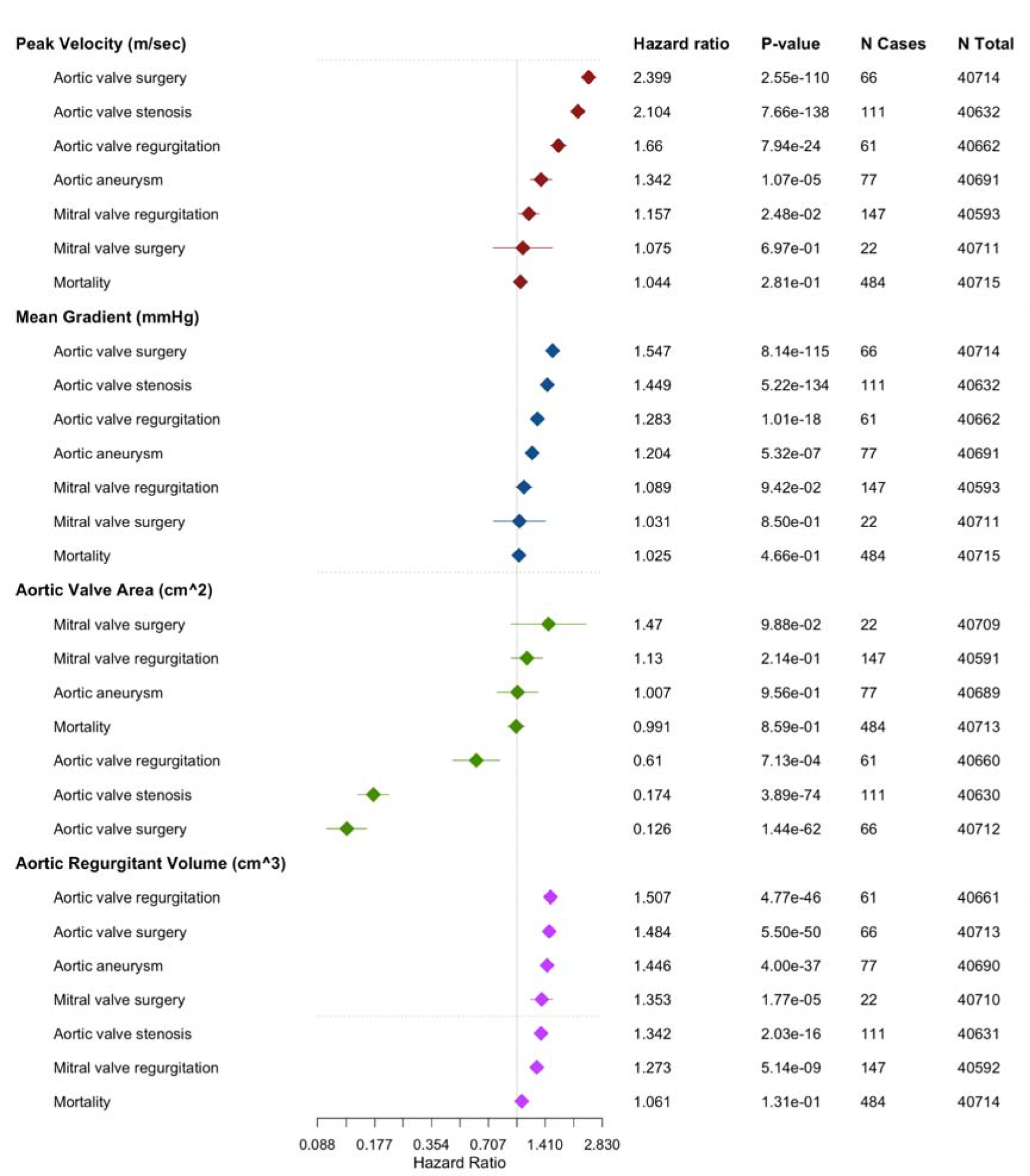
Incident valvular disease per standard deviation of aortic valve phenotypes. Cox hazard models of incident aortic and mitral valve disease per standard deviation of peak velocity, mean gradient, aortic valve area and aortic regurgitant volume. The models were adjusted for age and age^2^ at the time of MRI, the MRI serial number, sex, the first five principal components and the genotyping array.

For example, mitral regurgitant volume was a predictor of incident pulmonary hypertension (N=32, hazard ratio [HR] 1.8 per SD, P=1.5 × 10^−12^) and atrial fibrillation (N=517, HR 1.24 per SD, P=6.9 × 10^−09^). Mitral regurgitant volume was the strongest predictor of incident mitral valve surgery (N=19, HR 1.9 per SD, P=9.7 × 10^−24^) (**Supplementary Figure EE**). Over half of the participants who underwent mitral surgery were in the top 5% for mitral regurgitant volume (N=11, HR 22.6, P=7.5 × 10^−11^) (**Supplementary Table H**).

Aortic regurgitant volume was a predictor of aortic root procedures (N=12, HR 1.6 per SD, P=1.0 × 10^−24^) and non-root thoracic aortic procedures (N=15, HR 1.6 per SD, P=1.4 × 10^−29^), as well as heart failure (N=305, HR 1.18 per SD, P=9.2 × 10^−06^). When limited to participants with LVEF greater than or equal to 50% at the time of imaging, aortic regurgitant volume retained a similar effect estimate for future heart failure diagnosis (N=207, HR 1.19 per SD, P=1.0 × 10^−04^).

Participants with elevated peak velocity across the aortic valve were more likely to receive a diagnosis of CAD (N=748, HR 1.23 per SD of peak velocity, P=2.0 × 10^−14^) and heart failure (N=305, HR 1.23 per SD, P=2.4 × 10^−07^), and to undergo echocardiography (N=653, HR 1.3 per SD of peak velocity, P=5.4 × 10^−33^). Higher peak velocity was also associated with endocarditis (N=13, HR 1.8 per SD, P=2.3 × 10^−11^) and a non-statistically-significantly higher rate of cardiac arrest (N=38, HR 1.3 per SD, P=3.8 × 10^−3^). Those in the top 5% of peak velocity also had a non-statistically-significantly higher rate of all-cause mortality (N=49, HR 1.5, P=8.9 × 10^−3^) (**Supplementary Table H**).

### Risk of incident aortic valve surgery

Incident aortic valve surgery (N=66) was most strongly predicted by AVA (HR 8.0 per SD decrease, P=1.4 × 10^−62^), followed by peak velocity (HR 2.4 per SD increase, P=2.6 × 10^−110^) and the mean gradient (HR 1.5 per SD increase, P=8.1 × 10^−115^). We observed this relationship even after excluding those with evidence of at least moderate aortic stenosis (**Supplementary Figure FF**). Most of the risk accrued to those in the top 5% (bottom 5% for AVA) (**Supplementary Figure GG,** Figure 4): those participants had a hazard ratio between 90.6 (for peak velocity, N=57 out of 66 events were above the threshold, P=1.1 × 10^−35^) and 108.8 (for AVA, N=50 out of 66 events were below the threshold, P=1.4 × 10^−52^) compared to the remaining 95%. These findings were attenuated but remained significant after limiting the analysis to individuals without evidence of at least moderate aortic stenosis according to guideline criteria at the time of MRI: this left 33 incident cases of aortic valve surgery, of which 24 were in the top 5% for mean gradient (HR 36.5, P=9.7 × 10^−20^) and peak velocity (HR 36.8, P=7.7 × 10^−20^), and 17 were in the bottom 5% for AVA (HR 43.0, P=5.4 × 10^−23^) (**Supplementary Table H**).

**Figure 4:**
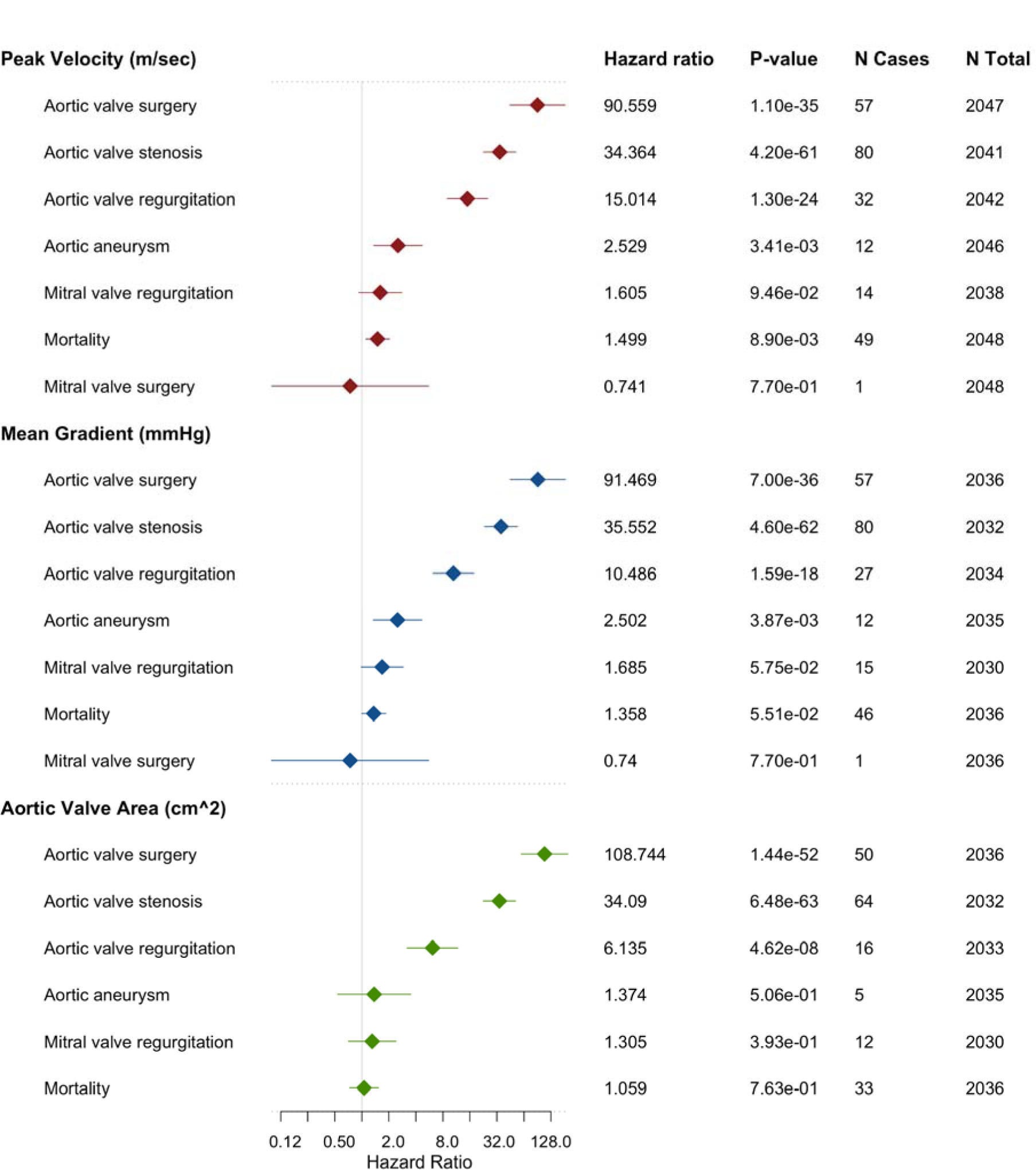
Stratified risk prediction for incident valvular disease. Cox hazard models of incident aortic and mitral valve disease in top 5% of participants vs remaining participants for peak velocity and mean gradient. Incident aortic valve disease of bottom 5% vs remaining participants for aortic valve area shown. The models were adjusted for age and age^2^ at the time of MRI, the MRI serial number, sex, the first five principal components and the genotyping array.

To obtain a qualitative perspective on the aortic velocity relationships, we then used linear models with spline terms to estimate the association between mean gradient, peak velocity, and AVA and the risk of undergoing valve surgery (**Supplementary Figure HH**). Participants with a mean gradient below 5mmHg had little risk of going on to have aortic valve surgery, compared to a nearly 35% chance of surgery near 20mmHg. In a *post hoc* analysis that grouped participants based on mean gradient (below 5mmHg [N=40,455], between 5-10mmHg [N=1,899], and above 10mmHg [N=170]), fewer than 0.5% of those with mean gradient below 5mmHg at the time of MRI had undergone valve surgery during follow-up time compared with nearly 1.5% with a mean gradient between 5-10mmHg. In contrast to this, nearly 40% of those with a mean gradient >10mmHg at the time of MRI had undergone valve surgery during mean 3.1 ± 2.0 years of follow-up time (**Supplementary Figure II**).

### Association with blood-based biomarkers

During the baseline visit, blood samples were collected from participants and used to measure 30 common biomarkers. We sought to assess whether these biomarkers were associated with the aortic flow-based phenotypes, which were ascertained a mean 9.6 years (SD 2.2 years) after enrollment (Figure 5 **and Supplementary Table I**). We observed that urate (0.078 SD increase per SD, P=6.1 × 10^−41^) and triglycerides (0.057 SD increase per SD, P=1.4 × 10^−30^) were associated with higher peak velocity. We also observed an association of apolipoprotein B (ApoB) (0.033 SD increase per SD, P=3.1 × 10^−12^) and lipoprotein a (Lp [a]) (0.027 SD increase per SD, P=4.8 × 10^−07^) with higher peak velocity. The strongest associations with mean gradient were found with urate (0.080 SD increase per SD, P=2.7 × 10^−30^) and sex hormone-binding globulin (SHBG)(−0.071 SD decrease per SD, P=9.2 × 10^−37^) but associations were also seen with C-reactive protein (0.056 SD increaser per SD, P=4.4 × 10^−32)^ and triglycerides (0.057 SD increase per SD, P=2.2 × 10^−31^).

**Figure 5:**
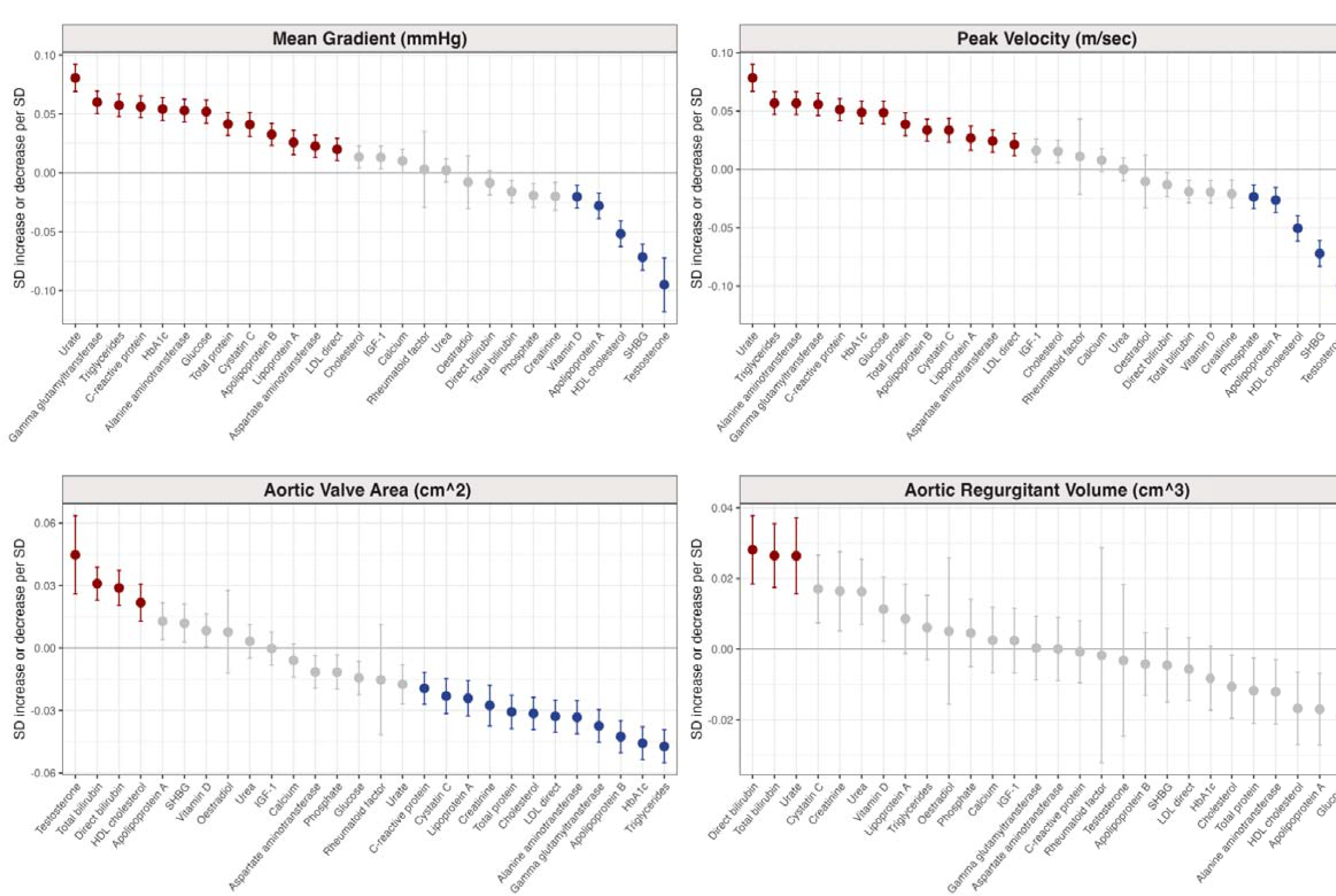
Association of biomarkers with aortic valve traits. Linear regressions of 30 biomarkers measurements ∼9.6 years before cardiac MRI and their association with mean gradient, peak velocity, aortic valve area and aortic regurgitant volume. Shown as standard deviation increase or decrease of aortic valve traits per standard deviation of biomarker. Red color indicates increased aortic valve trait per standard deviation and significant after Bonferroni correction for multiple testing. Blue color indicates decreased aortic valve trait per standard deviation and significant after Bonferroni correction for multiple testing. Grey indicates no significant association between biomarker and aortic valve trait after Bonferroni correction for multiple testing. The models were adjusted for the cubic spline of age at enrollment, the cubic spline of age at the time of MRI, the MRI serial number, sex, the first five principal components, and the genotyping array.

Moreover, strong associations with smaller AVA were observed with triglycerides (−0.047 SD decrease per SD, P=1.7 × 10^−31^) and HbA1c (−0.046 SD decrease per SD, P=5.3 × 10^−30^), followed by ApoB (−0.042 SD decrease per SD, P=4.1 × 10^−27^). C-reactive protein, a marker of inflammation, was associated with both smaller AVA (−0.019 SD decrease per SD, P=7.5 × 10^−07^) and higher peak velocity (0.051 SD increase per SD, P=5.9 × 10^−27^). Using the available NMR metabolomic data, we observed that higher levels of glycoprotein acetyls were associated with higher peak velocity (0.095 SD, P=6.1 × 10^−24^) and decreased AVA (−0.06 SD, P=6.2 × 10^−16^) (additional findings in **Supplementary results** and **Supplementary Table J**).

## Discussion

We used deep learning to study cardiac valvular function in nearly 50,000 UK Biobank participants. These measurements permitted a large-scale characterization of normal valve function and the relation between abnormalities of valve function, cardiovascular diseases, and their clinical and biological risk factors.

First, we described the normal ranges for aortic valve functional parameters within a large healthy sub-cohort of more than 30,000 healthy individuals. In all subgroups, the 95^th^ percentile cutoffs for AVA, peak velocity, and mean gradient did not exceed current thresholds defining moderate AS (1.5cm^2^, 3m/sec, and 20mmHg, respectively)^18,19^. Men and women had similar peak velocities and mean gradients, while AVA was larger for men than for women—even after indexing on body surface area. These observations align with sex-agnostic thresholds for gradient-based measurements in the assessment of AS, but also give credence to the notion that continuity equation-based AVA may warrant a sex-specific cutoff, as recently proposed ^20^. Whether adverse clinical outcomes are better explained by uniform or sex-specific thresholds—and at which boundary those thresholds should be drawn—will be important to understand through future efforts.

Second, within a predominantly healthy population sample, we noted that the annual decrement in AVA was minimal: 0.03cm^2^ per year. This stands in contrast to the accelerated progression documented in individuals exhibiting symptoms of AS, where severity appears to correlate with the rate of progression. For instance, a recent systematic review and meta-analysis encompassing 5,450 patients from 24 studies reported annual AVA reductions of 0.07 cm², 0.08 cm², and 0.09 cm² for mild, moderate, and severe AS, respectively ^21^.

Third, we did observe that sub-threshold measurements were still markers for future risk. For example, after excluding participants with any measurement consistent with at least moderate aortic stenosis, participants in the top 5% of mean gradient measurements still had a 36.5-fold increased risk of undergoing aortic valve surgery during follow-up compared to all other participants, while those in the bottom 5% of AVA had a 43.0-fold increased risk.

The current EACVI/ASE consensus document defines distinct thresholds for moderate and severe disease using peak velocity, mean gradient, and AVA ^17^. It should be noted that these traits are not used in isolation clinically, as international guidelines recommend the assessment of all three for the diagnosis of AS ^18, 19^. We observed that the AVA cutoffs uniformly categorized participants into higher severity classes than the velocity and gradient-based cutoffs. It is also notable that only peak velocity has been given a lower bound defining mild disease in the consensus guidelines (2.6m/s), but not AVA or mean gradient. Previous studies focused on either smaller cohorts or selected patient groups ^22, 23^, but nevertheless showed that the likelihood of surviving without valve replacement gradually decreases with increasing peak velocity starting at >3m/s ^24^. We now extend those observations of progressive risk down to peak velocities as low as 1.75m/s, mean gradient measurements as modest as 5mmHg, and AVA as large as 1.75cm^2^. The values identified in this study, unbiased by confounding by imaging indication, may provide an updated estimate of where the boundaries between normal and mild aortic stenosis might be drawn. These thresholds may also inform future efforts for conducting presymptomatic population surveillance for early disease detection or assessing response to preventive therapies.

Fourth, our findings provide additional evidence linking inflammatory and lipoprotein biology to normal variation in aortic valve function. Prevalent diagnoses of hypercholesterolemia and CAD were associated with higher peak velocity, greater mean gradient, and smaller AVA. So, too, were higher quantitative measurements of triglycerides, ApoB, and Lp(a) taken at UK Biobank enrollment (an average of 9.6 years before imaging). In addition to lipoproteins, we observed associations with inflammatory biomarkers such as C-reactive protein and glycoprotein acetyls. Notably, these relationships were not confined to those with evidence of subclinical moderate (or greater) AS; rather, the associations had similar magnitudes after excluding such participants. Both lipids and inflammation have long been linked to AS and its endophenotypes ^25^. For example, tissue analysis of explanted stenotic aortic valves has revealed inflammatory remodeling in lipid-rich valvular tissue, linking both lipid-formation and inflammation to AS ^26^. Genetic studies have previously implicated inflammation and lipid biology in AS, with associations at both the interleukin-6 locus and the *LPA* locus ^27, 28^. However, randomized controlled trials of lipid lowering therapy with statins have been neutral in preventing progression of AS ^6, 7^. The present findings suggest that the relationship between elevated lipoproteins and aortic valve dysfunction may begin well before the onset of overt clinical disease. This observational evidence raises the question of whether there is a role for early intervention among high-risk individuals, analogous to the concept of lifetime lipoprotein exposure’s impact on CAD risk. Currently, several approaches to lowering Lp(a) levels are undergoing clinical trials ^29, 30^. These trials may provide clearer causal evidence on the relationship between lipoprotein biology and AS.

Fifth, the data permitted large-scale observations of mitral regurgitation. Although likely underdiagnosed in this study population compared to prior population estimates^31^ due in part to heterogeneity of symptoms ^32^, participants with known mitral valve prolapse had significantly elevated mitral regurgitant volumes, providing technical affirmation of our approach to inferring mitral regurgitant volume from aortic and left ventricular stroke volumes. Mitral valve abnormalities are common in HCM ^33^, and here we observed that participants with HCM also tended to have elevated mitral regurgitant volumes. In turn, mitral regurgitant volume was predictive of ultimately undergoing mitral valve surgery as well as being predictive of both pulmonary hypertension and atrial fibrillation. These findings fit with a pathophysiological model whereby regurgitant mitral flow can elevate left atrial pressure, impairing left atrial compliance, predisposing to atrial fibrillation, and contributing to elevated pulmonary artery pressures and right ventricular afterload.

In summary, we performed large-scale derivation of left-sided valvular function by using a deep learning model to assist with extracting flow-encoded measurements. We estimated normal reference ranges and identified phenotypic and disease associations. We anticipate that exploration of causal mechanisms influencing valvular function will be a valuable path forward for future efforts.

### Limitations

Our study should be viewed in the context of its several limitations. It is based on the UK Biobank which is a primarily European-ancestry cohort that is healthier than the general population in the United Kingdom^34^. The MRI cohort tends to be healthier than the overall UK Biobank due to immortal time between enrollment and imaging; the follow-up period after imaging is short, with a small number of incident disease events.

Because of the velocity-encoding acquisition mode, any flow that was not perpendicular to the plane of imaging would be underestimated. Flow imaging at the level of the left ventricular outflow tract was unavailable, precluding the calculation of AVA by the standard continuity equation. Because most images were acquired distal to the level of the aortic valve, planimetry could not be accurately assessed; for the same reason, excluding participants with morphologically bicuspid valves could not be done with confidence, although bicuspid valve is expected to be present in 1% of the population. Further, the observations made with respect to continuity-based AVA may not be concordant with valve area as measured by planimetry. Because of variation in the level of image acquisition, the average diameter fell in between those previously reported for the aortic root ^35^ and the ascending aorta ^11^. Because of the limited time resolution of MRI (here, 30 images per cardiac cycle), mean gradient may have greater variance compared to transthoracic echocardiography. For epidemiological analyses, disease assignment was based on ICD and procedural codes, which are lagging indicators of symptoms and disease onset. Several diagnoses—including bicuspid valve disease and mitral valve prolapse—appeared to be predicted by aortic and mitral flow patterns, but were likely present at the time of MRI and not yet clinically diagnosed. An assessment of valvular disease-related symptoms at the time of MRI was unavailable. Finally, the deep learning models are not expected to generalize to other datasets without fine tuning to local MRI acquisition protocols.

## Online Methods

### Study overview

This project was conducted under UK Biobank application #41664. It was approved by the MGB institutional review board (IRB; protocol 2019P003144) and considered exempt by the UCSF IRB (#22-37715). In brief, UK Biobank participants with cardiac MRI were analyzed to calculate physical measurements from velocity-encoded data. The epidemiology of these phenotypes and their relationship with prevalent and incident disease were analyzed.

### Magnetic resonance imaging in UK Biobank

The UK Biobank is a prospective, general population-based cohort study that enrolled ∼500,000 individuals in the UK between the ages 40-69 years from 2006-2010 ^12^. Informed consent was obtained from all participants. Comprehensive phenotyping including questionnaires about family history, physical traits, life-style factors, laboratory values and imaging was obtained for each participant. Inpatient electronic health records from Hospital Episode Statistics (England), Patient Episode Database (Wales) and Scottish Morbidity Records (Scotland) as well as National Health Service death registries are linked to the cohort ^36^.

The imaging substudy of the UK Biobank is planned to perform 1.5 Tesla cardiac MRI in ca. 100,000 participants with ca. 50,000 studies in individual participants available as of the time of manuscript preparation ^37^. The cardiac MRI images were captured in a 20 minute study using a Siemens 1.5 Tesla MAGNETOM Aera scanner (Siemens Healthineers, Erlangen, Germany). In this study, phase contrast flow images aimed to be placed above the sinotubular junction at end-diastole were used. Phase contrast imaging uses magnitude scans as reference scans and velocity-encoded scans to create phase contrast velocity maps with a planned standard velocity encoding (VENC) of 2 m/s ^38^. Over the cardiac cycle, 30 images with a slice thickness (depth) of 6 mm and a voxel size of 1.77 x 1.77 mm with retrospective gating were acquired. Consequently, the amount of time represented by each image varied among participants (due to varying heart rates).

### Statistical analysis

Statistical analyses were performed with R 4.2.2 unless otherwise stated. Phenotype distributions and correlation grids were plotted using R 4.2.2 with the *ggplot2* and *ggcorrplot* packages. Extreme values beyond 5 standard deviations above or below the mean were removed from the phenotype distribution plots. Association testing between each phenotype and age and sex was performed with the *lm* function in the R 4.2.2 base package. Association testing for repeated measures was performed with the *lm* function, forcing a zero intercept. Visualization of the fraction of the population undergoing valve surgery for a given mean gradient was estimated with a generalized additive model using a 7-degree-of-freedom natural spline on the mean gradient.

### Generating training and test data for segmentation

One cardiology fellow (S.K.) manually annotated pixels in 950 randomly selected images from the CINE sequences within the UK Biobank imaging series “flow_XXX_tp_AoV_bh_ePAT” within the UK Biobank (where XXX represents the velocity encoding parameter in centimeters per second). Another cardiologist (J.P.P.) manually annotated pixels of the ascending aorta in 30 randomly selected images. For image annotation and model training, only the CINE channel (and not the magnitude channel or the velocity encoding channel) was used.

A cumulative density function (CDF) was generated from the non-background, non-lung components of the manually annotated CINE images as defined by the manually traced segmentation masks. The pixel intensities were rescaled based on the inverse of this cumulative density function, similar to the approaches described by Nyúl, et al ^39^, and Shinohara, et al ^40^. Any pixel value below the lowest rescaled value was set to 0 and any pixel value above the highest rescaled value was set to 1; all other values within the CDF’s range were linearly mapped between 0 and 1 (**Supplementary Table A**). This was applied to all CINE images (but not the velocity-encoding images) from all participants as a preprocessing step before applying the deep learning algorithm both during training and during model inference.

### Segmentation with deep learning

The modeling procedure is similar to that described previously ^11, 41^. A U-Net based deep learning model from the fastai v2.7.9 library was constructed in PyTorch v.1.12 ^16^ using a ResNet34 encoder that had been pre-trained with natural images from ImageNet ^42, 43^. The model was trained on 800 of the manually traced segmentation masks and 150 additional samples were used for validation.

All images were scaled to be in the range of 0-1 using the CDF matching described above. The images were normalized with the following constants: mean 0.298 and standard deviation 0.288. During training, several affine augmentations of the inputs were applied with probability 99%, including: affine rotation from 0-360 degrees; translation of up to 10% from the center; zooming between 90-110% of the original scale; shear between −10 and 10; and reflection padding. Brightness was varied by up to by 20%. Random erasing was permitted 25% of the time, in which between 2-10% of the image was replaced with background in rectangular blocks. In all cases, any geometric transformation applied to the input image was also applied to the segmentation mask to preserve the mask’s correspondence with the input image. During validation and testing, only scaling from 0-1 and normalization were applied, without other augmentations.

The model was fully unfrozen and trained for 500 epochs with PyTorch using the AdamW optimizer with the default weight decay (0.01). The training schedule was a OneCycleLR schedule, which was described by Smith and Topin for model superconvergence ^44^. With the one-cycle training schedule, the scheduler was allowed to achieve a maximum learning rate of 1e-4. The loss function was a focal loss with a gamma parameter of 2 ^45^. At each epoch, a Dice score was computed for both the ascending and descending aortic blood pools in the validation samples and averaged; if this average value was superior to any prior epoch, then the model weights were saved. The model weights from the epoch with the best validation Dice score (epoch 309) were saved for downstream use. The model was then applied to all available CINE images.

### Segmentation quality control

After applying the model to all CINE images, the output segmentation masks underwent heuristic quality control for the ascending aortic blood pool using an approach that has been previously described ^11^. Images without exactly one connected component for the labeled structure were flagged. The instantaneous frame-to-frame change in the number of pixels attributed to the labeled structure was computed, and any study above or below 5 standard deviations from the mean shift was flagged. Images were also excluded if both systolic and diastolic phases were not detected. Any flagged image was removed from analysis. Only participants with complete studies (those having 30 images that satisfied quality control) were retained for downstream analysis.

### Extracting velocity-encoded values

Velocity at each pixel was computed within the aortic blood pool. The CINE segmentation masks were overlaid on their paired velocity-encoded images, which allowed for velocity-based measurements to be computed for labeled regions. For each aortic blood pool pixel at each of 30 time points throughout the cardiac cycle, the through-plane velocity at that pixel was extracted from the paired velocity-encoded image. For each image, the VENC value was retrieved from the Siemens header (DICOM group 0×0029, element 0×1010). “Bits_stored’’ was uniformly defined as 12 in the DICOM metadata (DICOM group 0×0028, element 0×0101). Therefore, the pixel data encoded a range of intensity values from 0 through 4095 (i.e., 2^12^−1). These were remapped to velocity values with units of centimeters per second, ranging from -VENC to +VENC, using Formula 1.

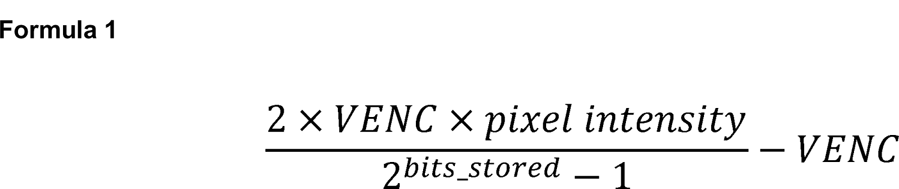

That formula yielded a through-plane velocity value for each pixel. Aggregating these values from all pixels at each frame allowed for the calculation of bulk properties such as Velocity Time Integral (VTI; necessary for computing the AVA) and forward stroke volume.

### Definition of systole

We created a heuristic to identify systole for each participant. We first computed the mean pixel velocity in the ascending aorta at all 30 frames, identifying the frame with the greatest mean velocity. Systole was defined to be the phase starting from the first image in the sequence with a mean velocity that shared the same sign as the greatest mean velocity. Systole was defined to continue until at least the frame with the greatest mean velocity; subsequent frames were checked for two conditions: first, if the mean velocity had a sign flip, systole was terminated. Second, if the prior frame had a mean velocity less than 20% that of the greatest mean velocity, and the current frame had a greater mean velocity than the prior frame, systole was terminated. Diastole was considered to begin on the frame after the termination of systole, and to continue until the frame before the start of systole. (As an example, if systole was found to begin in frame 3 and end in frame 10, then diastole was assigned to be from frame 11-30 as well as frames 1-2.) Velocity-derived measurements were then computed separately within systole and diastole for the ascending aorta.

### Deriving phenotypes from segmentation and velocity encoding data

Eight phenotypes with physical units were derived: ascending aortic diameter (cm), greatest average velocity (cm/sec), peak velocity (cm/sec), mean peak gradient (“mean gradient”; mmHg), forward stroke volume (mL), AVA (cm^2^), mitral regurgitant volume (mL), and aortic regurgitant volume (mL).

Aortic diameter was computed from the CINE segmentation masks, after accounting for the physical representation of each pixel in centimeters from the DICOM metadata and computing the elliptical minor axis diameter at its largest point in systole using image moments, as previously described ^11, 46^.

Peak velocity was determined by identifying the 99th percentile velocity for all pixels at each time point, and retaining the maximum value at any time point during systole. The 99th percentile velocity was selected as a heuristic to reduce spuriously high peak velocity values attributable to noise (Figure 2). The 99th percentile velocity was used to compute the gradient across the valve (using the simplified Bernoulli equation 4v^2^) for each frame, and then the mean value of that gradient at all time points during systole was taken as the mean gradient. Greatest average velocity—which is not used clinically—was calculated by computing the mean velocity of all pixels in the aortic blood pool at each time point during systole, and then retaining the maximum of those values.

Forward stroke volume was computed by summing all pixel-wise forward volumes (computed by multiplying velocity, width, height, and duration of each frame) for the aortic blood pool during systole.

Aortic valve area was computed by dividing forward stroke volume by VTI ^47^. The 99th percentile pixel velocity was treated as the boundary of the VTI envelope throughout the cardiac cycle in order to calculate VTI for this formula.

Aortic regurgitant volume was computed by summing all per-pixel retrograde volumes during any time point in the diastole where the net aortic stroke volume was retrograde.

Left ventricular volumes were required to produce estimates of the mitral regurgitant volume. We used the volumes calculated by the unmodified model from Pirruccello et al ^41^. Mitral regurgitant volume was then computed by subtracting the forward aortic stroke volume from the left ventricular stroke volume. Mitral regurgitant volumes that were estimated to be negative were set to missing.

### Aortic phenotypes in individuals without evidence of at least moderate aortic stenosis

For peak velocity, mean gradient and aortic valve area we created a “bounded” phenotype that excluded any individuals with evidence for at least moderate AS if one or more of the following conditions were met: peak velocity > 3m/sec, mean gradient >20mmHg, AVA >1.5 cm^2^. This was done to assess downstream analyses without any influence of moderate-to-severe stenotic aortic valve disease. Moderate or greater disease was chosen because while guidelines provide measurement boundaries to distinguish mild from moderate aortic stenosis, most guidelines do not provide measurements that define a boundary between mild aortic stenosis and normal.

### Definitions of diseases and outcomes in UK Biobank

Definitions of diseases and outcomes are provided in **Supplementary Table B**. Generally, we used self-reported data as well as ICD codes (ICD-9, ICD-10), and procedural codes (OPCS-3, OPCS-4) from National Health Service registries and the inpatient data from Hospital Episode Statistics. Follow-up time was censored on September 30, 2021.

### Reference ranges

To calculate reference ranges, participants with aortic stenosis, coronary artery disease, heart failure, or hypertension were excluded. Participants without any of those diagnoses were included, even if their measurements suggested the likely presence of clinically undetected disease (e.g., AVA < 1.5cm^2^). Phenotype distributions were calculated by sex within four age bands (<55, 55-64, 65-74, >=75). Mean values ± standard deviation, as well as median values with 5% and 95% cutoffs, were reported. Then, reference ranges were defined separately for men and women. For each phenotype, five zones were defined: abnormally low (below the 5% cutoff in all age groups), borderline low (below the 5% cutoff in at least one age group), normal (between the 5% and 95% cutoff in all age groups), borderline high (above the 95% cutoff in at least one age group), and abnormally high (above the 95% cutoff in all age groups).

### Association with prevalent cardiovascular disease

Diseases already diagnosed at the time of imaging were tested for association with MRI-derived phenotypes. Participants with a diagnosis occurring after imaging were included, but labeled as not having disease for this analysis. The linear model was constructed such that the MRI-derived phenotype was the dependent variable, with disease as the independent variable of interest, with covariates including the MRI serial number, age and age^2^ at the time of MRI, sex, the first five genetic principal components of ancestry precomputed by the UK Biobank centrally, and the genotyping array ^36^. Phenotypes were scaled so that the effect estimates were in units of a 1 standard deviation (SD) change, to allow comparability across phenotypes.

### Association with incident cardiovascular disease

MRI-derived phenotypes were tested for association with incident disease diagnosed subsequent to imaging. Follow-up time was censored at the time of loss-to-follow-up or death where relevant, and after September 30, 2021 for remaining participants. We evaluated incident cardiovascular disease using Cox proportional hazard models using time from the date of imaging until the date of diagnosis or censoring as the outcome. We repeated this step using the top decile of distribution for each phenotype in a stratified analysis.

The Cox models were adjusted for age and age^2^ at the time of MRI, the MRI serial number, sex, the first five principal components and the genotyping array. The effect sizes were reported as hazard ratio of disease risk per standard deviation of the phenotype. Diseases with fewer than 10 incident events were excluded from the analysis.

### Stratified risk prediction for incident cardiovascular disease

To better understand the relationship of the MRI-derived phenotypes in the tails of distribution, we performed a stratified analysis comparing the top 5% and in a separate analysis the bottom 5% of the participants with the remaining cohort for incident cardiovascular disease. We employed the same Cox hazard models as in the overall analysis and adjusted for age and age^2^ at the time of MRI, the MRI serial number, sex, the first five principal components and the genotyping array.

### Prediction of risk for future aortic valve surgery

To investigate the relationship of aortic valve traits with risk for future aortic valve surgery, we used splines to generate nonlinear models of the absolute value of either mean gradient, peak velocity or AVA to predict the cumulative risk for aortic valve surgery over the follow-up time to produce a qualitative visualization. This was performed in R, where a generalized additive model using a 7-degree-of-freedom natural spline on each phenotype was generated using the *ggplot2*::*geom_smooth* function.

For mean gradient, the participants were then stratified into three categories in *post hoc* fashion (<5mmHg, 5-10 mmHg and >10mmHg, respectively) to calculate the cumulative incidence (1-Kaplan Meyer estimate of the survival function) of undergoing aortic valve replacement.

### Continuous trait analysis of NMR metabolites and biomarkers

Linear regression was used to assess the relationship between each imaging-derived phenotype and biomarkers and nuclear magnetic resonance (NMR)-based metabolites in the UK Biobank. The blood-based measurements were taken at the time of enrollment, so the model was structured with the imaging-derived phenotype as the dependent variable and the biomarker as the independent variable, adjusting for the cubic spline of age at enrollment, the cubic spline of age at the time of MRI, the MRI serial number, sex, the first five principal components, and the genotyping array. Both imaging-derived phenotypes and biomarkers were scaled to a mean of zero and a standard deviation of one to facilitate cross-comparison. Therefore, the effect sizes reported represent the coefficient per standard deviation of dependent variable per standard deviation change in the independent variable.

## Data Availability

The derived phenotypes from application #41664 will be returned to the UK Biobank. Access to UK Biobank data may be requested by researchers in academic, commercial, and charitable organizations.

## Acknowledgements

Dr. Kany was supported by the Walter Benjamin Fellowship from the Deutsche Forschungsgemeinschaft (521832260). Dr. Rämö was supported by a research fellowship from the Sigrid Jusélius Foundation. Dr. Nauffal is supported by NIH grant 5T32HL007604-35. Dr. Khurshid is supported by NIH grant T32HL007208. This work was supported by grants from the National Institutes of Health to Dr. Lau (K23-HL159243), Dr. Ellinor (11R01HL092577, 1R01HL157635, 5R01HL139731), Dr. Ho (R01HL134893, R01HL140224, R01HL160003, K24HL153669), and Dr. Pirruccello (K08HL159346). This work was also supported by grants to Dr. Lau from the American Heart Association (853922), Dr. Ellinor from the American Heart Association Strategically Focused Research Networks (18SFRN34110082), and from the European Union (MAESTRIA 965286). Dr. Lindsay is supported by the Toomey Fund for Aortic Dissection Research and by NIH grant (1R01NS125353).

## *UK Biobank* acknowledgment

UK Biobank is generously supported by its founding funders the Wellcome Trust and UK Medical Research Council, as well as the British Heart Foundation, Cancer Research UK, Department of Health, Northwest Regional Development Agency and Scottish Government.

## References

1. Yadgir, S. et al. Global, Regional, and National Burden of Calcific Aortic Valve and Degenerative Mitral Valve Diseases, 1990–2017. Circulation 141, 1670–1680 (2020).

2. Pellikka, P. A. et al. Outcome of 622 adults with asymptomatic, hemodynamically significant aortic stenosis during prolonged follow-up. Circulation 111, 3290–3295 (2005).

3. Strange, G. et al. Poor Long-Term Survival in Patients With Moderate Aortic Stenosis. J. Am. Coll. Cardiol. 74, 1851–1863 (2019).

4. Ito, S. et al. Prognostic Risk Stratification of Patients with Moderate Aortic Stenosis. J. Am. Soc. Echocardiogr. 34, 248–256 (2021).

5. Lee, W. et al. Long-term Prognosis of Mild to Moderate Aortic Stenosis and Coronary Artery Disease. Journal of Korean Medical Science vol. 36 Preprint at https://doi.org/10.3346/jkms.2021.36.e47 (2021).

6. Cowell, S. J. et al. A randomized trial of intensive lipid-lowering therapy in calcific aortic stenosis. N. Engl. J. Med. 352, 2389–2397 (2005).

7. Chan, K. L. et al. Effect of Lipid lowering with rosuvastatin on progression of aortic stenosis: results of the aortic stenosis progression observation: measuring effects of rosuvastatin (ASTRONOMER) trial. Circulation 121, 306–314 (2010).

8. Otto, C. M. et al. Determination of the stenotic aortic valve area in adults using Doppler echocardiography. J. Am. Coll. Cardiol. 7, 509–517 (1986).

9. Cawley, P. J. et al. Prospective Comparison of Valve Regurgitation Quantitation by Cardiac Magnetic Resonance Imaging and Transthoracic Echocardiography. Circulation: Cardiovascular Imaging vol. 6 48–57 Preprint at https://doi.org/10.1161/circimaging.112.975623 (2013).

10. Defrance, C. et al. Evaluation of aortic valve stenosis using cardiovascular magnetic resonance: comparison of an original semiautomated analysis of phase-contrast cardiovascular magnetic resonance with Doppler echocardiography. Circ. Cardiovasc. Imaging 5, 604–612 (2012).

11. Pirruccello, J. P. et al. Deep learning enables genetic analysis of the human thoracic aorta. Nat. Genet. 54, 40–51 (2022).

12. Sudlow, C. et al. UK biobank: an open access resource for identifying the causes of a wide range of complex diseases of middle and old age. PLoS Med. 12, e1001779 (2015).

13. Pirruccello, J. P. et al. Development of a Prediction Model for Ascending Aortic Diameter Among Asymptomatic Individuals. JAMA 328, 1935–1944 (2022).

14. Gomes, B. et al. Genetic architecture of cardiac dynamic flow volumes. bioRxiv (2022) doi:10.1101/2022.10.05.22280733.

15. Córdova-Palomera, A. et al. Cardiac imaging of aortic valve area from 34 287 UK Biobank participants reveals novel genetic associations and shared genetic comorbidity with multiple disease phenotypes. Circ. Genom. Precis. Med. 13, e003014 (2020).

16. Paszke, A., et al. PyTorch: An imperative style, high-performance deep learning library. arXiv [cs.LG] (2019).

17. Baumgartner, H., Chair et al. Recommendations on the echocardiographic assessment of aortic valve stenosis: a focused update from the European Association of Cardiovascular Imaging and the American Society of Echocardiography. Eur. Heart J. Cardiovasc. Imaging 18, 254–275 (2017).

18. Writing Committee Members et al. 2020 ACC/AHA guideline for the management of patients with valvular heart disease: A report of the American college of cardiology/American heart association joint committee on clinical practice guidelines. J. Am. Coll. Cardiol. 77, e25–e197 (2021).

19. Vahanian, A. et al. 2021 ESC/EACTS Guidelines for the management of valvular heart disease: developed by the Task Force for the management of valvular heart disease of the European Society of Cardiology (ESC) and the European Association for Cardio-Thoracic Surgery (EACTS). Eur. Heart J. 43, 561–632 (2022).

20. Masiero, G. et al. SexlJSpecific Considerations in Degenerative Aortic Stenosis for FemalelJTailored Transfemoral Aortic Valve Implantation Management. J. Am. Heart Assoc. 11, e025944 (2022).

21. Willner, N. et al. Aortic Stenosis Progression: A Systematic Review and Meta-Analysis. JACC Cardiovasc. Imaging 16, 314–328 (2023).

22. Rosenhek, R. et al. Mild and moderate aortic stenosis. Natural history and risk stratification by echocardiography. Eur. Heart J. 25, 199–205 (2004).

23. Eveborn, G. W., Schirmer, H., Heggelund, G., Lunde, P. & Rasmussen, K. The evolving epidemiology of valvular aortic stenosis. the Tromsø study. Heart 99, 396–400 (2013).

24. Otto, C. M. et al. Prospective study of asymptomatic valvular aortic stenosis. Clinical, echocardiographic, and exercise predictors of outcome. Circulation 95, 2262–2270 (1997).

25. Pérez de Isla, L., et al. Lipoprotein (a), LDL-cholesterol, and hypertension: predictors of the need for aortic valve replacement in familial hypercholesterolaemia. Eur. Heart J. 42, 2201–2211 (2021).

26. Mohty, D. et al. Association between plasma LDL particle size, valvular accumulation of oxidized LDL, and inflammation in patients with aortic stenosis. Arterioscler. Thromb. Vasc. Biol. 28, 187–193 (2008).

27. Thériault, S. et al. Genetic Association Analyses Highlight IL6, ALPL, and NAV1 As 3 New Susceptibility Genes Underlying Calcific Aortic Valve Stenosis. Circ Genom Precis Med 12, e002617 (2019).

28. Small, A. M. et al. Multiancestry Genome-Wide Association Study of Aortic Stenosis Identifies Multiple Novel Loci in the Million Veteran Program. Circulation (2023) doi:10.1161/CIRCULATIONAHA.122.061451.

29. Tsimikas, S. et al. Lipoprotein(a) reduction in persons with cardiovascular disease. N. Engl. J. Med. 382, 244–255 (2020).

30. Yeang, C. et al. Effect of pelacarsen on lipoprotein(a) cholesterol and corrected low-density lipoprotein cholesterol. J. Am. Coll. Cardiol. 79, 1035–1046 (2022).

31. Freed, L. A. et al. Prevalence and clinical outcome of mitral-valve prolapse. N. Engl. J. Med. 341, 1– 7 (1999).

32. Avierinos, J. F., Gersh, B. J. & Melton, L. J. Natural history of asymptomatic mitral valve prolapse in the community. ACC Current Journal Review vol. 12 37–38 Preprint at https://doi.org/10.1016/s1062-1458(02)01020-6 (2003).

33. Klues, H. G., Maron, B. J., Dollar, A. L. & Roberts, W. C. Diversity of structural mitral valve alterations in hypertrophic cardiomyopathy. Circulation 85, 1651–1660 (1992).

34. Fry, A. et al. Comparison of Sociodemographic and Health-Related Characteristics of UK Biobank Participants With Those of the General Population. American Journal of Epidemiology vol. 186 1026–1034 Preprint at https://doi.org/10.1093/aje/kwx246 (2017).

35. Nekoui, M. et al. Spatially Distinct Genetic Determinants of Aortic Dimensions Influence Risks of Aneurysm and Stenosis. J. Am. Coll. Cardiol. 80, 486–497 (2022).

36. Bycroft, C. et al. The UK Biobank resource with deep phenotyping and genomic data. Nature 562, 203–209 (2018).

37. Raisi-Estabragh, Z., Harvey, N. C., Neubauer, S. & Petersen, S. E. Cardiovascular magnetic resonance imaging in the UK Biobank: a major international health research resource. Eur. Heart J. Cardiovasc. Imaging 22, 251–258 (2021).

38. Petersen, S. E. et al. UK Biobank’s cardiovascular magnetic resonance protocol. J. Cardiovasc. Magn. Reson. 18, (2015).

39. Nyul, L. G., Udupa, J. K. & Zhang, X. New variants of a method of MRI scale standardization. IEEE Transactions on Medical Imaging vol. 19 143–150 Preprint at https://doi.org/10.1109/42.836373 (2000).

40. Shinohara, R. T. et al. Statistical normalization techniques for magnetic resonance imaging. Neuroimage Clin 6, 9–19 (2014).

41. Pirruccello, J. P. et al. Genetic analysis of right heart structure and function in 40,000 people. Nat. Genet. 54, 792–803 (2022).

42. He, K., Zhang, X., Ren, S. & Sun, J. Deep residual learning for image recognition. arXiv [cs.CV] 770–778 (2015).

43. Krizhevsky, A., Sutskever, I. & Hinton, G. E. ImageNet classification with deep convolutional neural networks. Commun. ACM 60, 84–90 (2017).

44. Smith, L. N. & Topin, N. Super-convergence: very fast training of neural networks using large learning rates. in Artificial Intelligence and Machine Learning for Multi-Domain Operations Applications vol. 11006 369–386 (SPIE, 2019).

45. Lin, T.-Y., Goyal, P., Girshick, R., He, K. & Dollar, P. Focal Loss for Dense Object Detection. IEEE Trans. Pattern Anal. Mach. Intell. 42, 318–327 (2020).

46. Horn, B., Klaus, B. & Horn, P. Robot Vision. (MIT Press, 1986).

47. Yap, S.-C. et al. A simplified continuity equation approach to the quantification of stenotic bicuspid aortic valves using velocity-encoded cardiovascular magnetic resonance. J. Cardiovasc. Magn. Reson. 9, 899–906 (2007).

