## Supplementary Notes for "Assessment of valvular function in over 47,000 people using deep learning-based flow measurements"

^1^Cardiovascular Disease Initiative, Broad Institute of MIT and Harvard, Cambridge, Massachusetts, USA, ^2^Department of Cardiology, University Heart and Vascular Center Hamburg-Eppendorf, Hamburg, Germany, ^3^Institute for Molecular Medicine Finland (FIMM), Helsinki Institute of Life Science (HiLIFE), University of Helsinki, Helsinki, Finland, ^4^University of Minnesota Medical School, Minneapolis, Minnesota, USA, ^5^Department of Experimental Cardiology, Amsterdam UMC, University of Amsterdam, Amsterdam, Netherlands, ^6^Division of Cardiovascular Medicine, Brigham and Women's Hospital, Boston, Massachusetts, USA, ^7^Cardiology Division, Massachusetts General Hospital, Boston, Massachusetts, USA, ^8^Bakar Computation Health Sciences Institute, University of California San Francisco, San Francisco, California, USA, ^9^Cardiology Division, Beth Israel Deaconess Medical Center, Boston, Massachusetts, USA, ^10^Division of Cardiology, University of California San Francisco, San Francisco, California, USA, ^11^Harvard Medical School, Boston, Massachusetts, USA, ^12^Cardiovascular Research Center, Massachusetts General Hospital, Boston, Massachusetts, USA, ^13^Thoracic Aortic Center, Massachusetts General Hospital, Boston, Massachusetts, USA, ^14^Institute for Human Genetics, University of California San Francisco, San Francisco, California, USA

### Supplementary Results

#### Defining aortic valve phenotypes without evidence for aortic stenosis

To understand the relationships between aortic valve phenotypes and cardiovascular disease in patients without evidence for aortic stenosis at the time of MRI, we applied the typical boundary conditions defining moderate or greater aortic stenosis: peak velocity >3m/sec, mean gradient >20mmHg, AVA >1.5 cm^2^. For analyses throughout this document referring to “bounded” aortic valve phenotypes, those participants with MRI evidence for moderate (or greater) AS, or a prior history of aortic valve surgery, were excluded.

#### Prevalent cardiovascular disease in participants without evidence for aortic valve stenosis

In the participants without evidence for AS, we still observed an association of prevalent hypertension (N=14409) with higher bounded peak velocity (0.30 SD increase, P=2.1 x 10^-192^), higher bounded mean gradient (0.31 SD, P=8.5 x 10^-179^) and smaller bounded AVA (N=14408, -0.06 SD, P=2.9 x 10^-13^) **(Supplementary Table F**). Similarly, a diagnosis of hypercholesterolemia (N=10162) was associated with a similar pattern of higher bounded peak velocity (0.17 SD increase, P=6.3 x 10^-46^), higher bounded mean gradient (0.18 SD, P=1.8 x 10^-50^) and smaller bounded AVA (N=10161, -0.15 SD, P=1.6 x 10^-51^). In these participants, we also observed a relationship of prevalent coronary artery disease (N=2153) with higher bounded peak velocity (0.17 SD increase, P=1.7 x 10^-14^), higher bounded mean gradient (0.20 SD, P=3.2 x 10^-19^) and smaller bounded AVA (-0.20 SD, P=1.3 x 10^-27^).

#### Incident cardiovascular disease in participants without evidence for aortic valve stenosis

In participants without evidence of at least moderate aortic stenosis by either peak velocity, mean gradient and AVA, we found that the bounded peak velocity was still strongly predictive of incident AS (N=76, HR 2.43, P=3.1 x 10^-59^) but also a incident aortic valve surgery (N=33, HR 2.65, P=3.0 x 10^-36^) **(Supplementary Table G**). Bounded mean gradient behaved similarly to bounded peak velocity and was predictive of AS (N=76, HR 1.80, P=2.9 x 10^-72^) and aortic valve surgery (N=33, HR 1.87, P=8.5 x 10^-43^). The statistical power for smaller bounded AVA to predict AS (N=76, HR 4.56, P=4.5 x 10^-23^) and incident aortic valve surgery (N=33, HR 4.15, P=1.4 x 10^-10^) was slightly lower but associated with bigger effect sizes.

#### Association of blood-based biomarkers with greatest average velocity and volume-based phenotypes

#### For greatest average velocity, we observed robust association of SHBG (-0.07 SD decrease per SD, P=2.3 × 10^-39^) decreasing and alanine aminotransferase (0.04 SD increase per SD, P=4.1 × 10^-19^) increasing greatest average velocity **(Supplementary Table I**). The association between blood-based biomarkers and mitral and aortic regurgitant volumes were less robust. However, total bilirubin and direct bilirubin for instance were associated with increased mitral regurgitant volume (direct bilirubin: 0.03 SD increase, P=3.4 × 10^-07^) and aortic regurgitant volume (direct bilirubin: 0.03 SD increase, P=1.1 × 10^-08^). Urate was associated with increased forward stroke volume (0.07 SD increase, P=1.7 × 10^-38^) and SHBG (-0.05 SD decrease, P=5.4 × 10^-23^) with decreased forward stroke volume.

#### Association of blood-based biomarkers with aortic valve phenotypes in those without aortic stenosis

We also investigated the association of biomarkers taken at baselines with aortic valve phenotypes in participants without evidence of at least moderate aortic stenosis. SHBG was robustly associated with reduced bounded mean gradient (-0.08 SD decrease per SD, P=4.2 x 10^-48^) and reduced bounded peak velocity (-0.07 SD decrease, P=1.4 x 10^-39^) **(Supplementary Table I**). Similar observations were seen for Urate and an association with increased bounded mean gradient (0.09 SD increase, P=1.4 x 10^-52^) and bounded peak velocity (0.08 SD increase, P=1.4 x 10^-41^). The strongest associations for bounded AVA were seen with triglycerides (-0.04 SD decrease, P=7.2 x 10^-28^), HbA1c (-0.04 SD decrease, P=3.8 x 10^-27^) and ApoB (-0.04 SD decrease, P=6.8 x 10^-26^).

#### Association of metabolomic measurements with aortic valve traits

Then, we assessed the association between lipid measurements (per standard deviation) at enrollment and aortic valve function and size at the time of MRI using the nuclear magnetic resonance (NMR) metabolomics available for 11,264 participants with cMRI data^1^. This was done since prior genetic studies suggested lipids may play a role in aortic valve disease ^2,3^. Triglycerides in small HDL were associated with a smaller AVA and an increase in peak velocity **(Supplementary Table J**). Very Large HDL components were associated with decreased peak velocity and increased AVA.

#### Association of metabolomic measurement with aortic valve traits in those without aortic stenosis

Glycoprotein acetyls were robustly associated with bounded mean gradient (0.11 SD increase, P=1.3 x 10^-29^), bounded peak velocity (0.01 SD increase, P=2.3 x 10^-25^) and bounded AVA (-0.06 SD decrease, P=3.9 x 10^-16^) **(Supplementary Table J**). We also observed strong effect sizes in the association between triglycerides in small HDL and bounded mean gradient (0.01 SD increase, P=7.9 x 10^-18^) and bounded peak velocity (0.08 SD increase, P=3.5 x 10^-15^). For bounded AVA we observed robust effects with monounsaturated fatty acids (-0.05 SD decrease, P=3.5 x 10^-12^) and triglycerides in medium LDL (-0.05 SD decrease, P=1.8 x 10^-11^).

#### Supplementary Discussion

We observed greatest average velocity to be far less predictive of incident AS (HR=1.30 per SD, P=4.0 x 10^-3^) than the standard clinical measurements of peak velocity (HR=2.11 per SD, P=7.7 x 10^-138^), mean gradient (HR=1.45 per SD, P=5.2 x 10^-134^), or smaller AVA (HR=5.8 per SD, P=3.9 x 10^-74^). Instead, lower greatest velocity more strongly predicted a diagnosis of thoracic aortic aneurysm (HR=6.2 per SD, P=2.1 x 10^-15^)—and did so to an even greater degree than larger diameter itself (HR=4.5 per SD, P=7.0 x 10^-57^). The bottom 5% of greatest average velocity were strongly predictive of thoracic aortic procedures not involving the aortic root (HR=29.2, P=6.0 x 10^-10^). This suggests that when the aortic valve does not have a pathological degree of stenosis, the bulk velocity of the aortic blood pool is (inversely) driven by the size of its conduit, rather than by the function of the valvular apparatus below—which is consistent with Bernoulli’s principle. We note that this greatest average velocity has sometimes been referred to as a peak velocity ^4^, although it is distinct from the concept of peak velocity as used in the aortic stenosis literature and throughout the present manuscript.

References:

1.	Ritchie, S. C. *et al.* Quality control and removal of technical variation of NMR metabolic biomarker data in ∼120,000 UK Biobank participants. Preprint at https://doi.org/[10.1101/2021.09.24.21264079](http://dx.doi.org/10.1101/2021.09.24.21264079).

2.	Nazarzadeh, M. *et al.* Plasma lipids and risk of aortic valve stenosis: a Mendelian randomization study. *Eur. Heart J.* **41**, 3913–3920 (2020).

3.	Thanassoulis, G. *et al.* Genetic associations with valvular calcification and aortic stenosis. *N. Engl. J. Med.* **368**, 503–512 (2013).

4.	Gomes, B. *et al.* Genetic architecture of cardiac dynamic flow volumes. *bioRxiv* (2022) doi:[10.1101/2022.10.05.22280733](http://dx.doi.org/10.1101/2022.10.05.22280733).

### Supplementary Figures

#### Supplementary Figure AA: Flow diagram

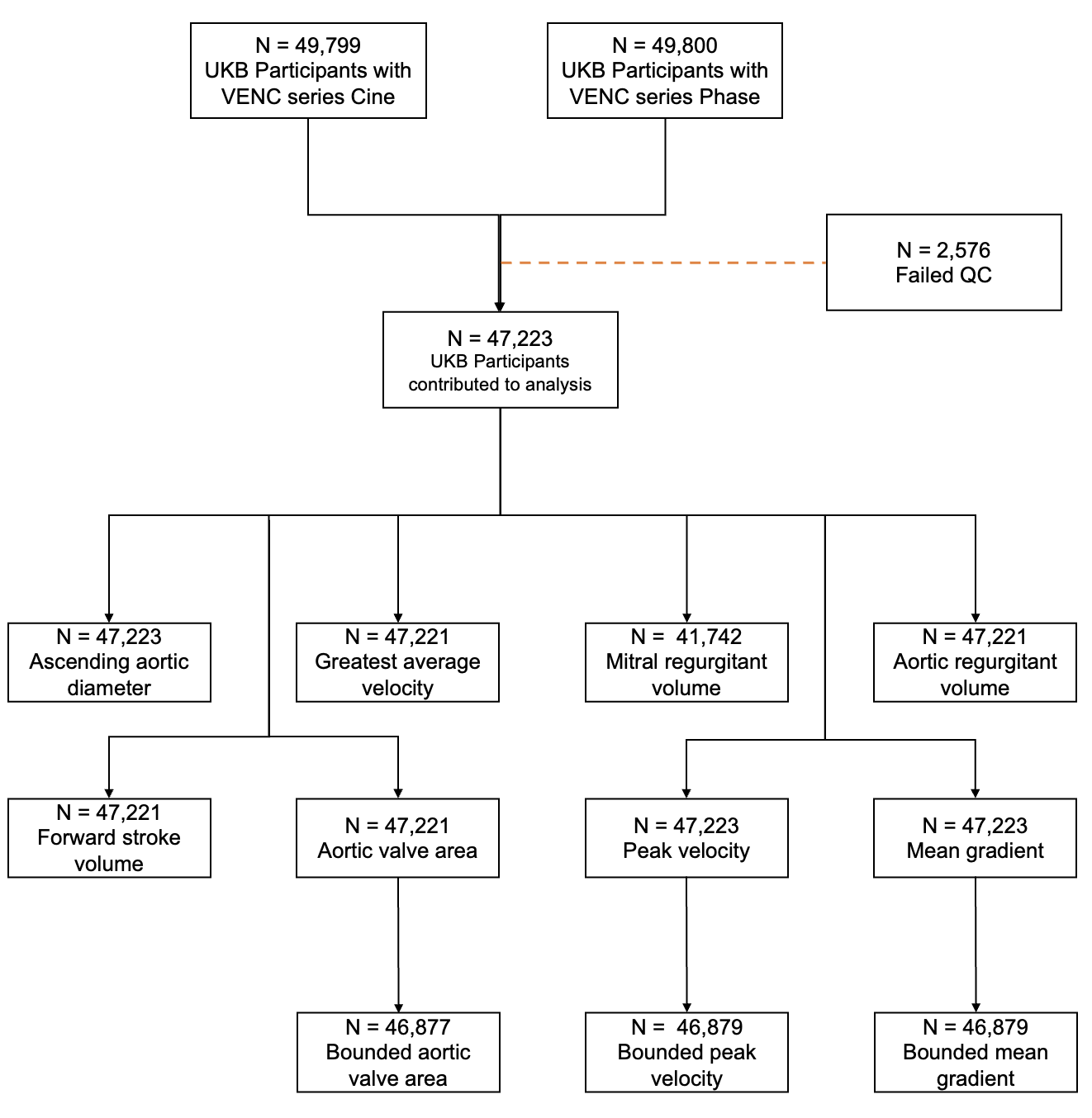

Consort flow chart of cohort construction showing the number of participants with available image series, the overall number of quality control failures and the number of participants that contributed to each respective phenotype.

#### Supplementary Figure BB: Phenotype distribution

**Panel A**

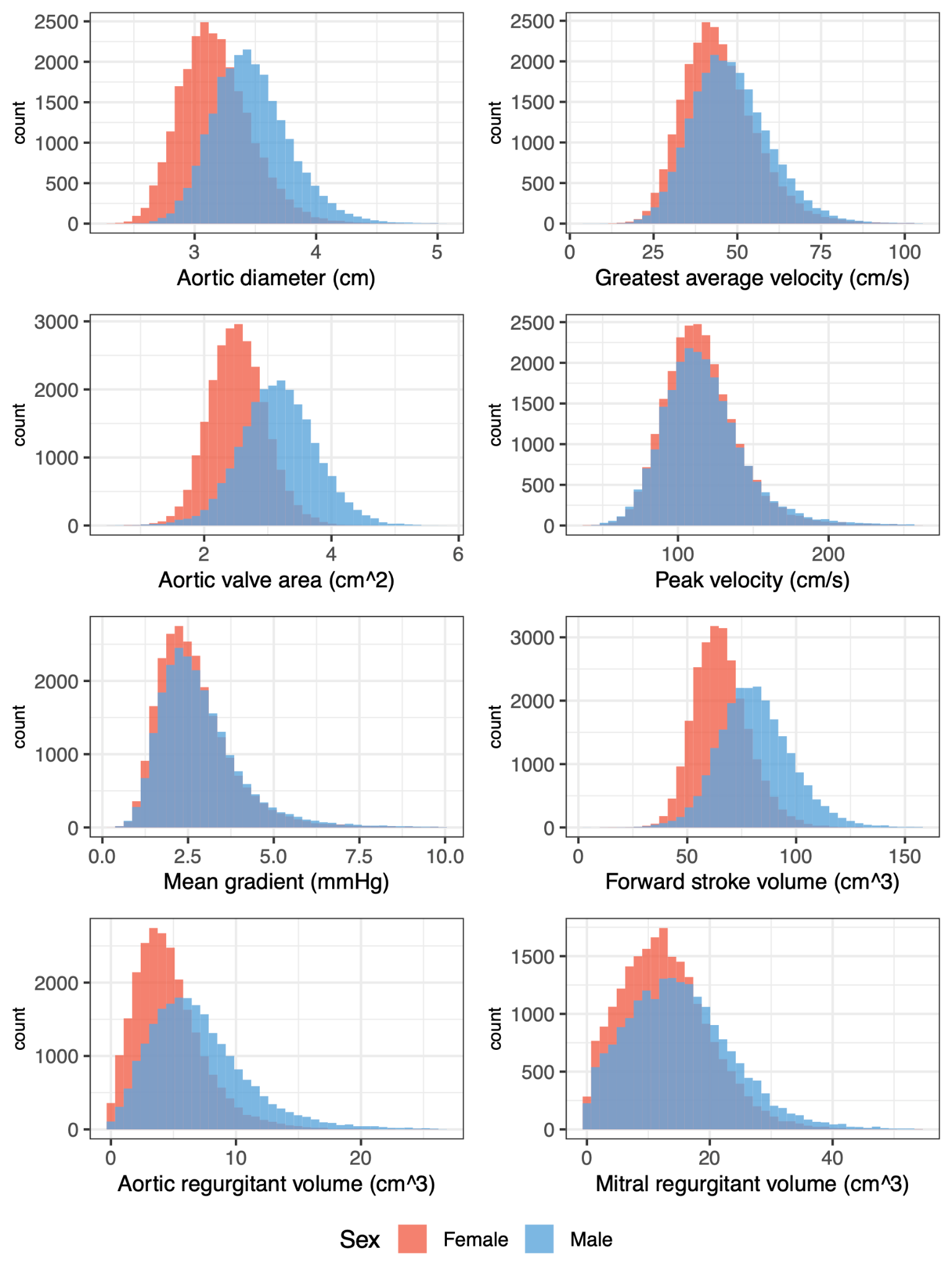

**Panel B**

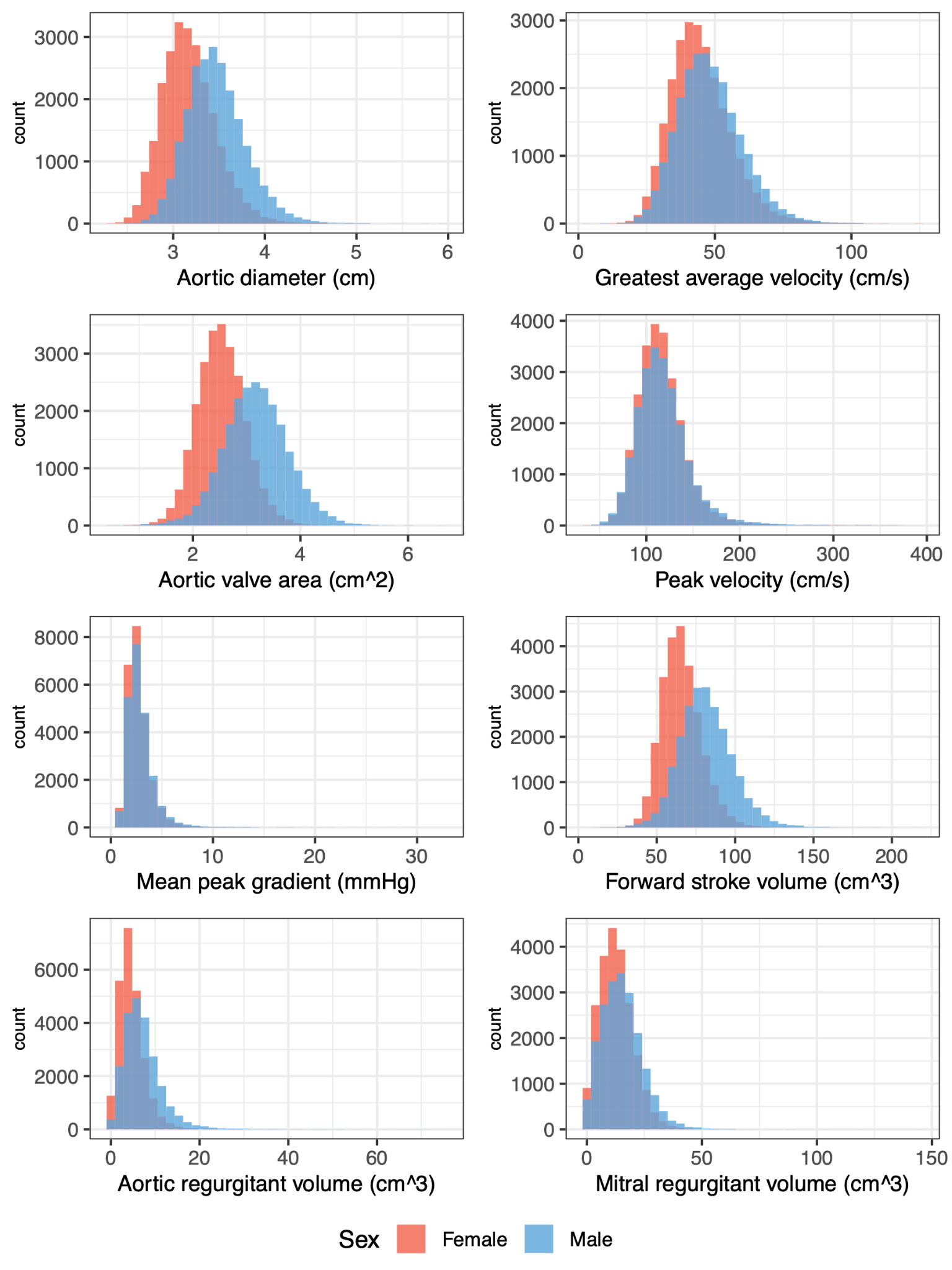

Histogram plots of the distribution of each phenotype by count and sex of the overall cohort included in the analysis. **Panel A**: samples with values beyond 5 standard deviations above the mean are removed to provide detail for the central distribution. **Panel B**: no limits are placed, in order to show the skew of the distributions driven by outliers.

#### Supplementary Figure CC: Correlation grid

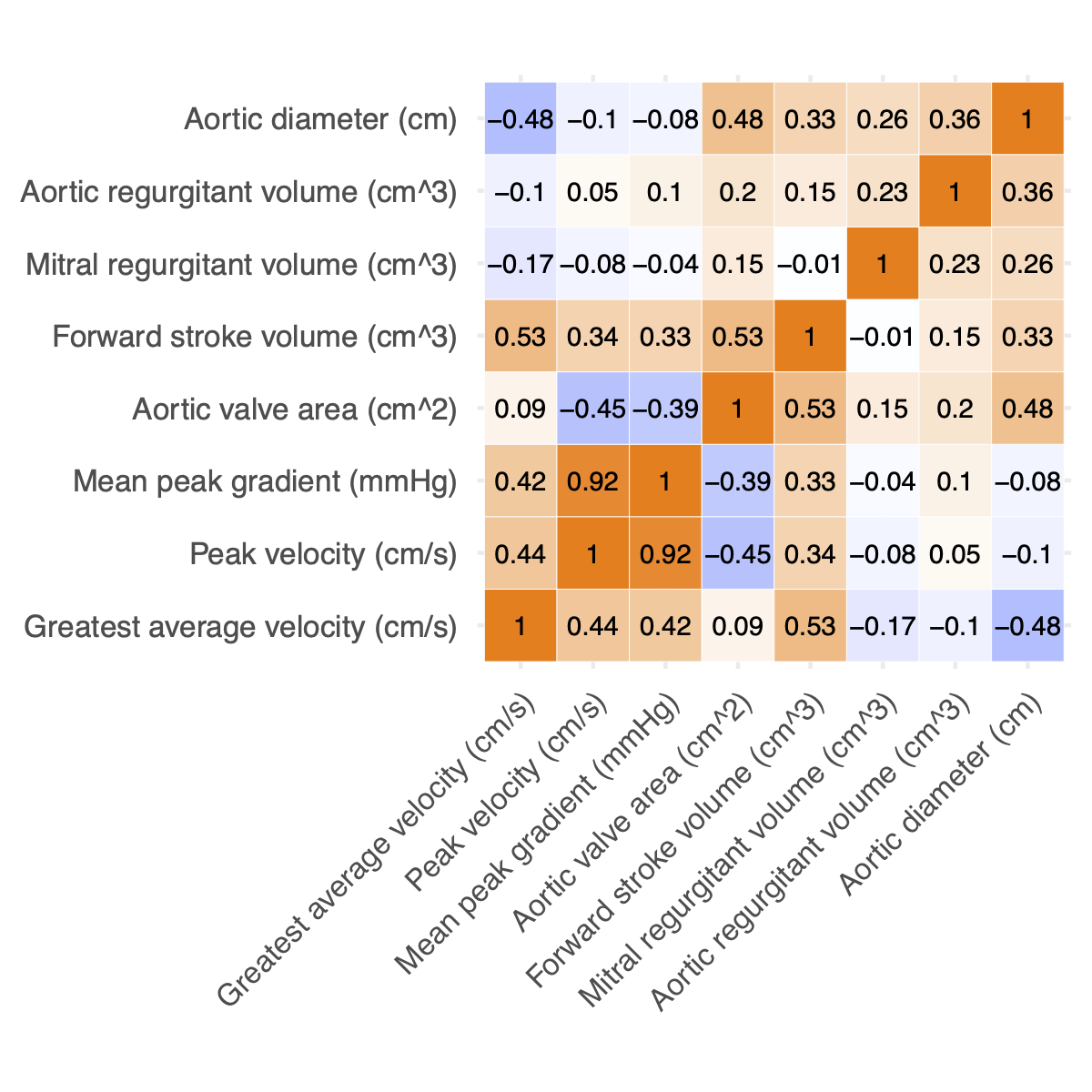

Correlation plots showing the correlation of each treat with other traits. Perfect correlation would be considered r=1 and perfect inverse correlation would be considered r=-1.

Supplementary Figure DD: Distribution of phenotypes and association with age and sex

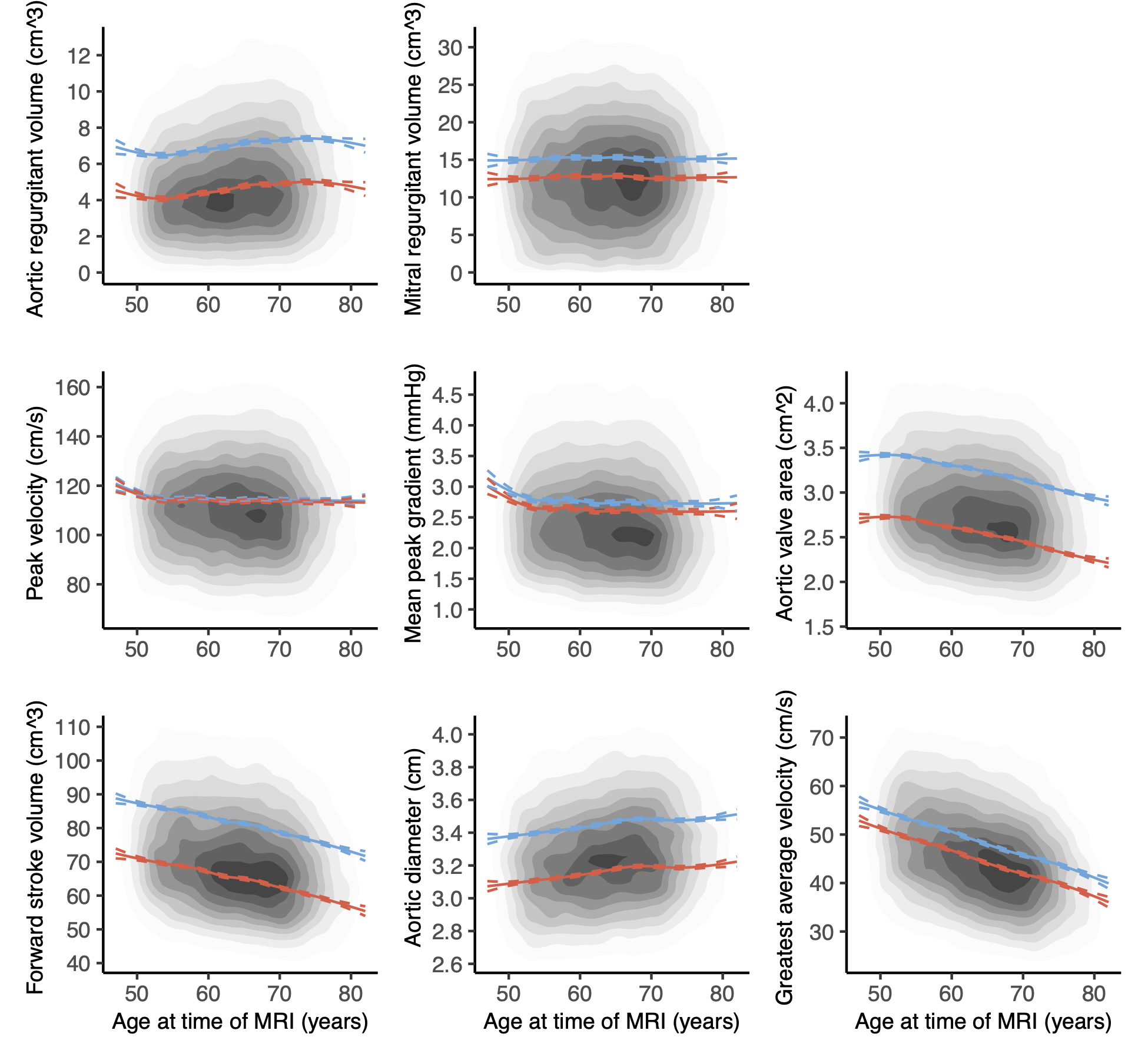

2D-density plots of flow-derived phenotypes by age for the healthy subset UK Biobank participants that underwent cardiac MRI (N=31,909). Using a 7-degree-of-freedom natural spline on age for each phenotype, the modeled average values for women are displayed in red and those for men are displayed in cerulean.

Supplementary Figure EE: Incident valvular disease with aortic diameter, maximum mean velocity, forward stroke volume and mitral regurgitant volume

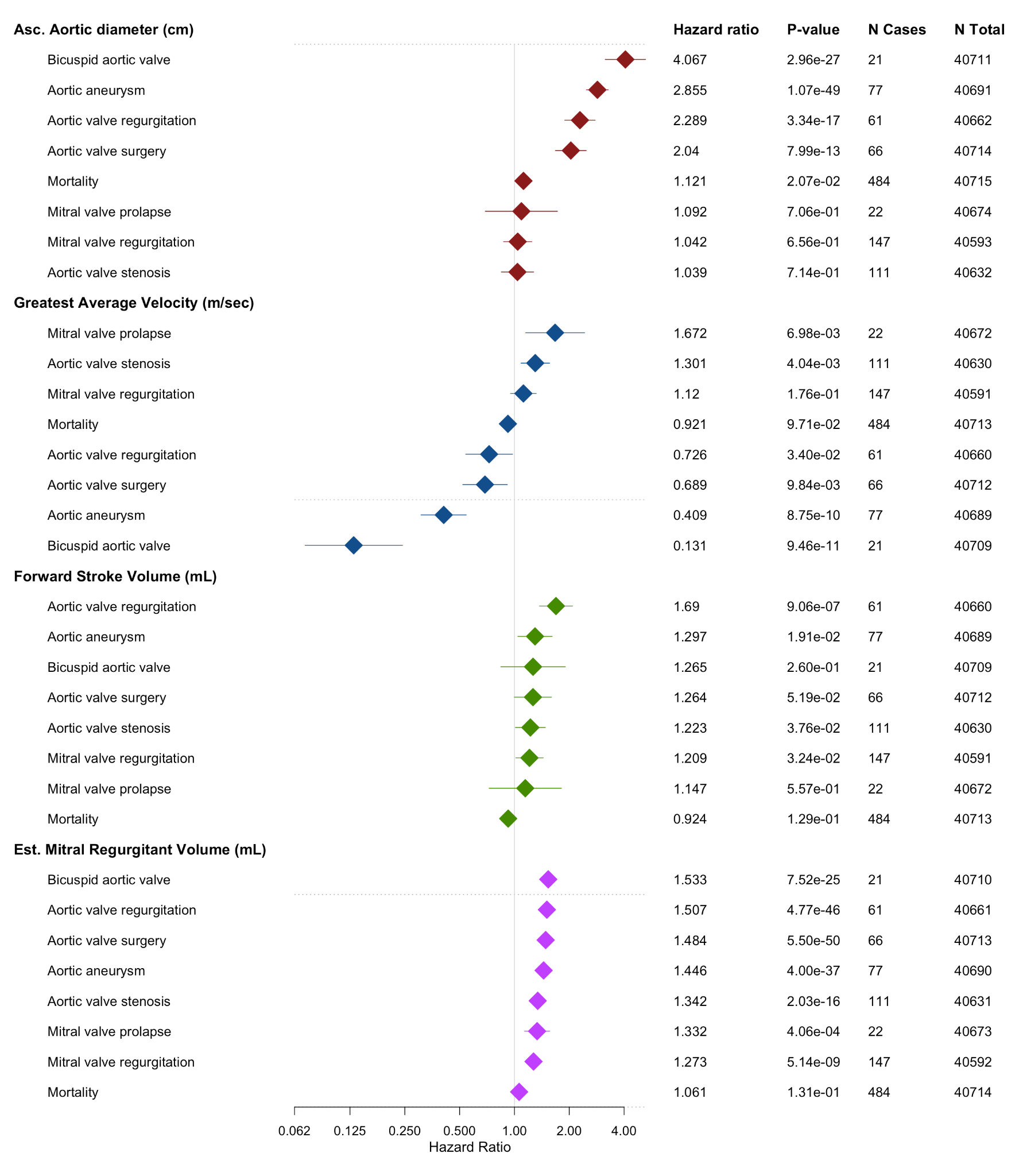

Cox hazard models of incident aortic and mitral valve disease per standard deviation of ascending aortic diameter, maximum mean velocity, forward stroke volume and mitral regurgitant volume.

##### Supplementary Figure FF: Incident valvular disease per standard deviation of bounded aortic valve phenotypes

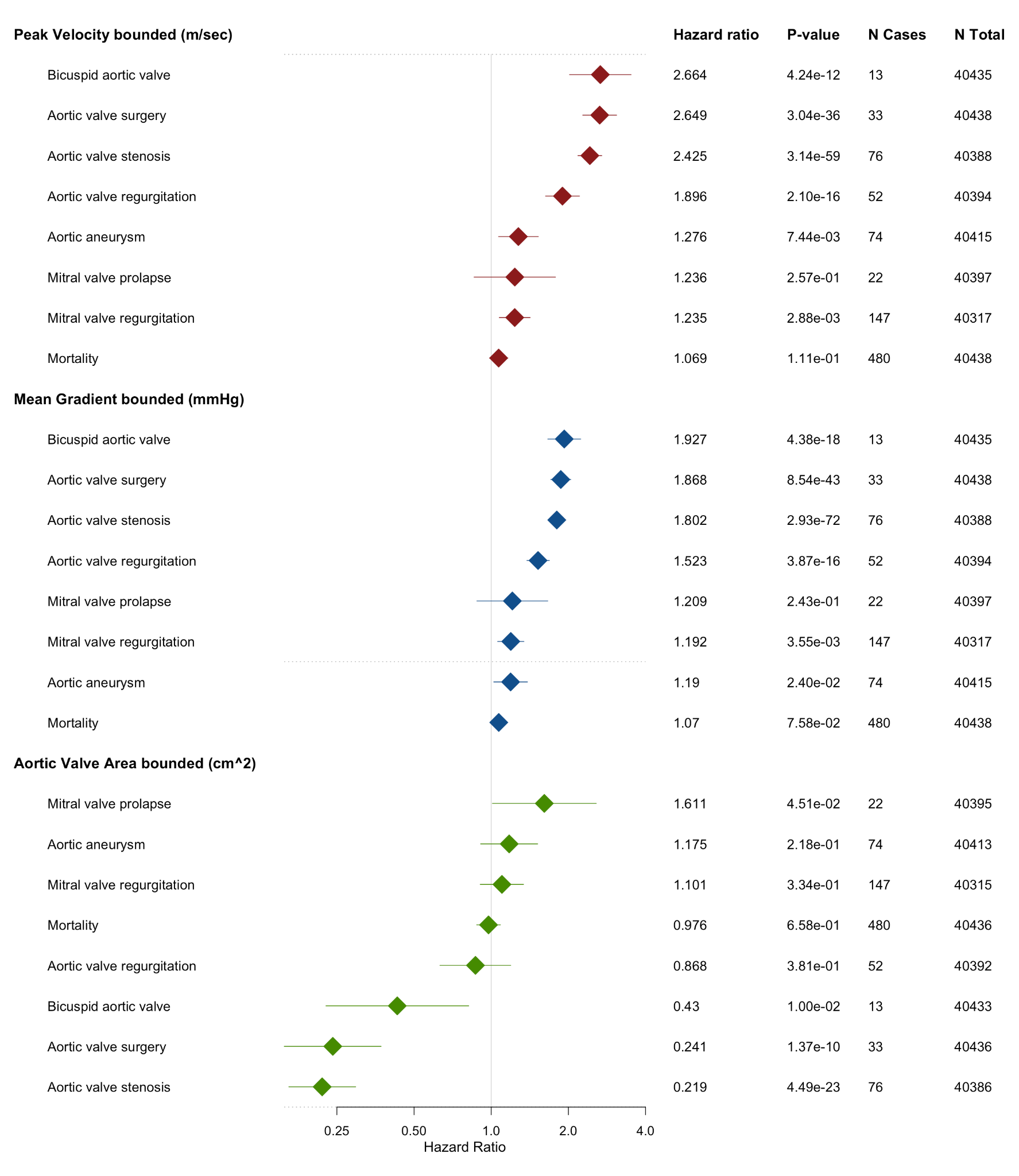

Cox hazard models of incident aortic and mitral valve disease per standard deviation of peak velocity, mean gradient and aortic valve area. Participants with evidence of at least moderate aortic stenosis (peak velocity >3m/sec, mean gradient >20mmHg, AVA >1.5 cm^2^) are excluded.

#### Supplementary Figure GG: Quantile groups of aortic valve phenotypes

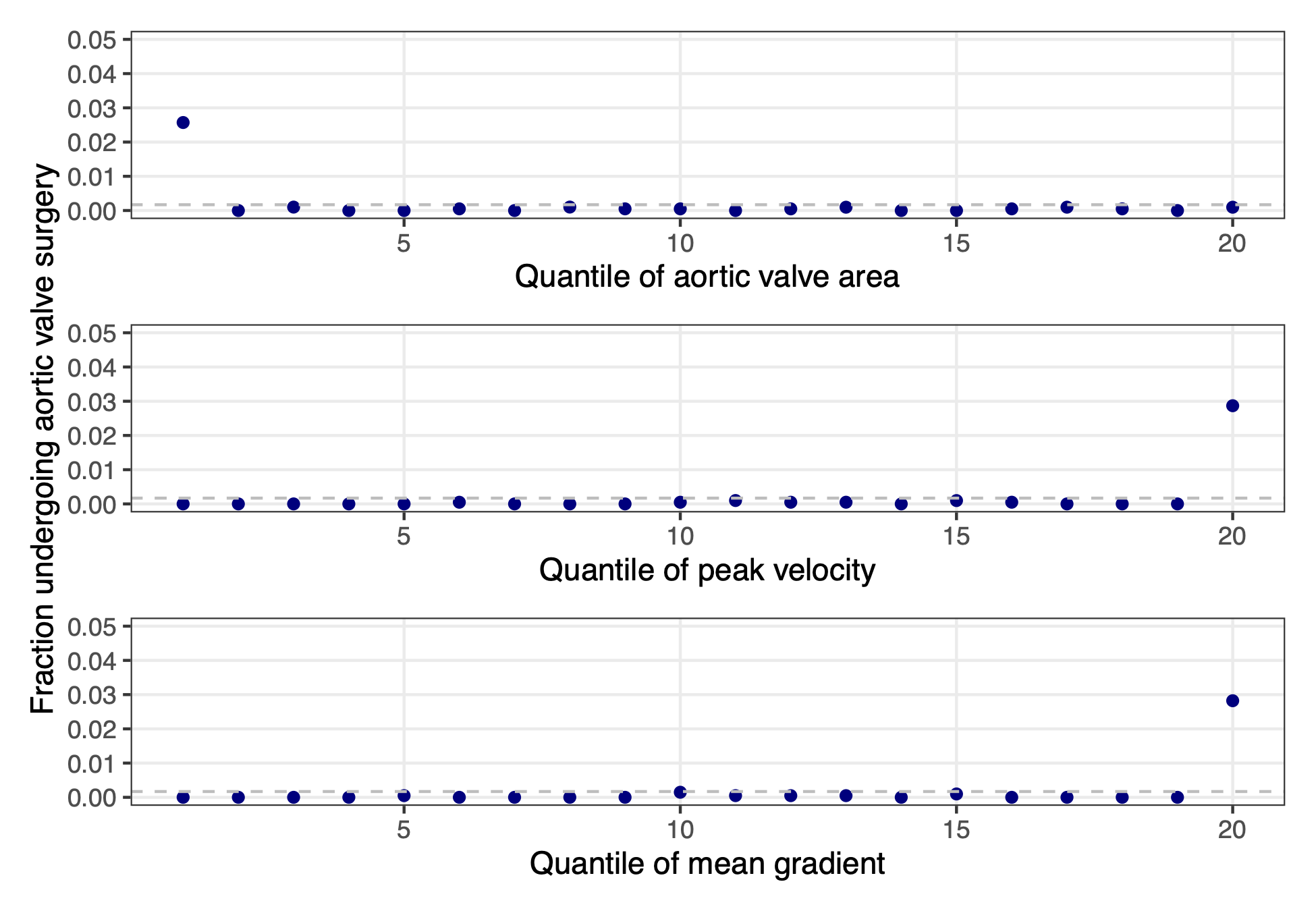

Each point represents 5% of the population, grouped based on phenotype values. Dashed lines represent the whole-population average.

#### Supplementary Figure HH: Relationship between mean gradient, peak velocity and aortic valve area and the risk of undergoing valve surgery in follow-up

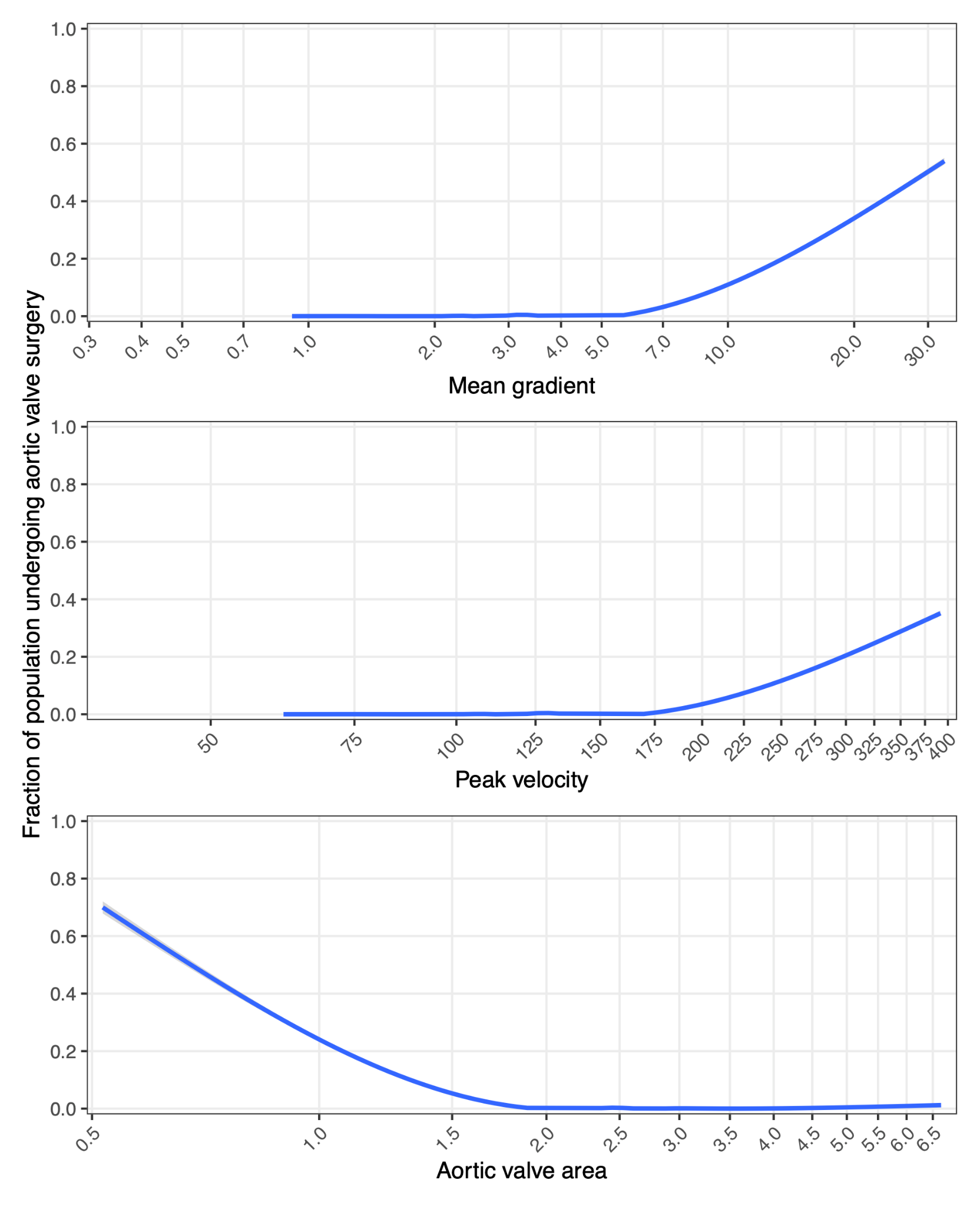

Nonlinear models of the fraction of population undergoing aortic valve surgery for the absolute values for each phenotype. The models were generalized additive models using a 7-degree-of-freedom natural spline on each phenotype.

#### Supplementary Figure II: KM Curves for mean gradient and aortic valve surgery

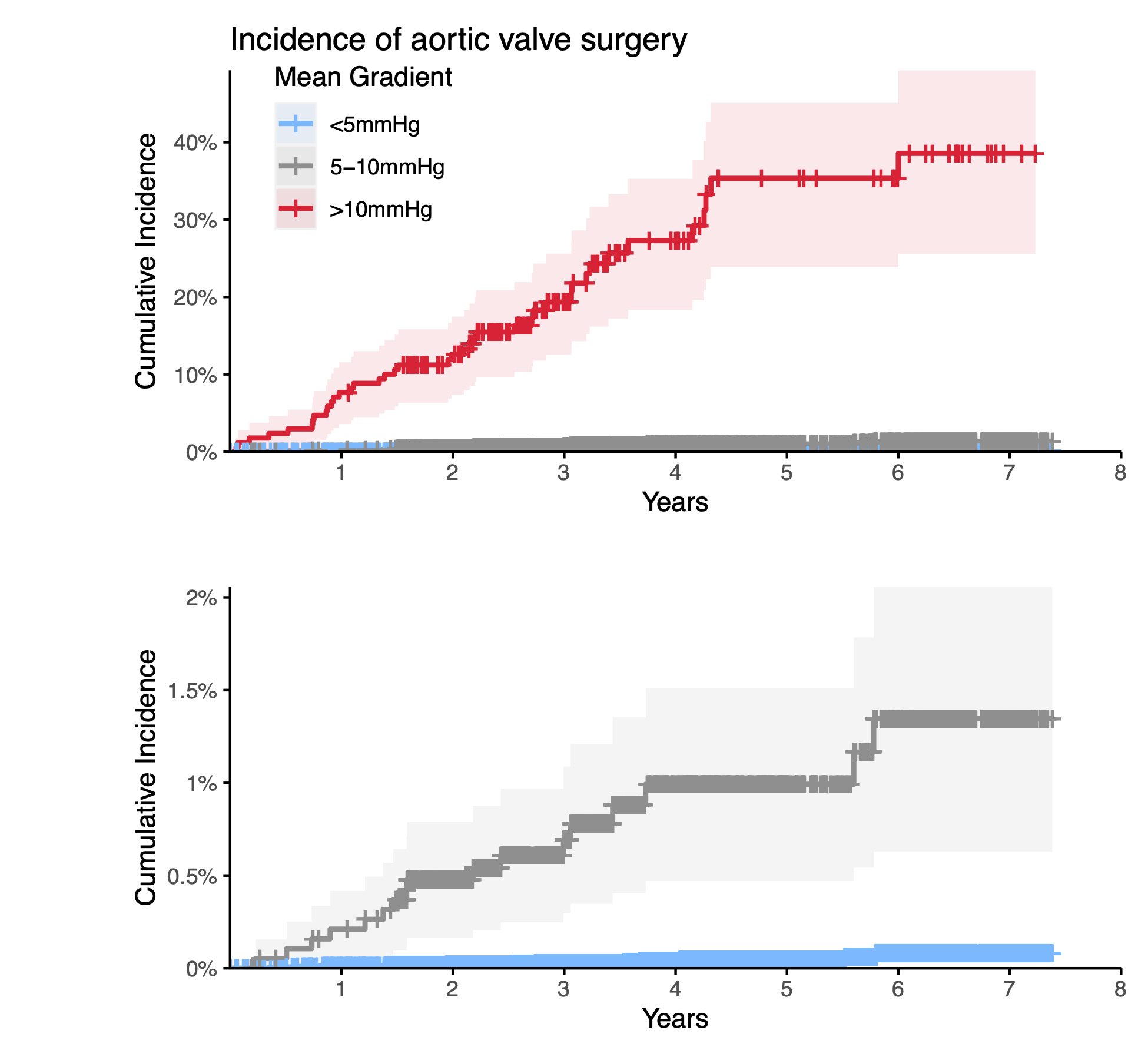

Top panel: cumulative incidence of aortic valve surgery based on stratified mean gradient groups (<5mmHg [N=40,455], 5-10mmHg [N=1,899], >10mmHg [N=170]). Bottom panel: cumulative incidence of aortic valve surgery with the top group (>10mmHg) excluded and incidence range narrowed to show the distinction between the lowest group and the middle group.
